# Tumor Associated Neutrophils Extend NETs To Spatially Drive Necroptosis In Glioblastoma

**DOI:** 10.64898/2026.10.25.26366446

**Authors:** Prasanna Vaddi, Yonghua Zhuang, Liqiong Liu, Steven Ingersoll, Schuyler Lee, Bifeng Gao, Adrie Van Bokhoven, Brent T. Harris, Sharon R. Pine, Hatim E. Sabaawy

**Affiliations:** Division of Medical Oncology, Department of Medicine, University of Colorado Denver, Anschutz Medical Campus; Aurora, CO, 80045, USA; Cancer Center Biostatistics and Bioinformatics Shared Resource, Department of Pediatrics, University of Colorado Denver, Anschutz Medical Campus; Aurora, CO, 80045, USA; Genomic Shared Resource, Department of Medicine, University of Colorado Denver, Anschutz Medical Campus; Aurora, CO, 80045, USA; Department of Pathology, University of Colorado Denver, Anschutz Medical Campus; Aurora, CO, 80045, USA; Pathology Shared Resource, Department of Pathology, University of Colorado Denver, Anschutz Medical Campus; Aurora, CO, 80045, USA; Department of Neurosurgery, University of Colorado Denver, Anschutz Medical Campus; Aurora, CO, 80045, USA; Department of Neurology, University of Colorado Denver, Anschutz Medical Campus; Aurora, CO, 80045, USA; University of Colorado Cancer Center; Aurora, CO, 80045, USA

**Keywords:** glioblastoma, pseudopalisading-necrosis, DAMP, Neutrophil exocellular traps (NETs), necroptosis

## Abstract

**Background:** Glioblastoma is pathologically characterized by extensive pseudopalisading necrosis, a hallmark linked to therapeutic resistance and dismal prognosis. While traditionally viewed as a passive ischemic event, the precise radiogenomic drivers and spatial immune architectures orchestrating active tumor necrosis remain poorly understood.

**Methods:** We integrated macroscopic clinical MRI radiomics with public genomic datasets (n=3,639), single-cell spatial transcriptomics, and high-resolution protein immunofluorescence (IF) assays in a novel co-mutant murine model to map the necrotic microenvironment.

**Results:** Radiomic analysis of patient MRIs demonstrated that high tumoral necrotic volume serves as a powerful independent biomarker for poor long-term survival. This macro-structural necrosis phenotype is radiogenomically driven by cell-intrinsic *TP53*, *PTEN*, and *NF1* (PPN) co-mutations. Spatial transcriptomic profiling of PPN-deficient tumors revealed distinct cellular topography, separating a proliferative, invasive niche (EGFR^+^ SOX^+^, Type^Inv^) from a hyper-aggressive necrotic niche (ERBB3^+^, Type^Nec^). Mechanistically, Type^Nec^ cells deploy a prominent CCL5/CXCL2 chemokine milieu that actively recruits and prompts tumor-associated neutrophils to undergo neutrophil extracellular trap (NET) formation. This overactive neutrophil infiltrate drives a feed-forward loop of intrinsic tumor necroptosis mediated by the release of the damage-associated molecular pattern (DAMP) protein HMGB2. This cascade could be therapeutically dismantled using targeted anti-inflammatory compounds. Finally, multi-cohort clinical validation confirmed that an HMGB2^high^ signature dictates a high-risk, myeloid-rich immunosuppressive microenvironment in glioblastoma patients.

**Conclusions:** Our findings redefine glioblastoma necrosis as an active, mutation-driven immunopathological process. The HMGB2-mediated DAMP necroptosis cascade and associated myeloid chemokines provide robust prognostic biomarkers and immediate therapeutic vulnerabilities to disrupt the immunosuppressive necrotic niche.

*Key Points:*

- **Clinical Stratification via Radiomics:** Preoperative MRI analysis of 1,079 glioblastoma patients establishes a necrotic core volume threshold of 18.57% as a robust, non-invasive biomarker for early survival divergence and dismal long-term prognosis.
- **Genomic Orchestration of Necrosis:** Cross-species modeling identifies a high-risk somatic PPN co-mutation profile yielding two topographically distinct niches: an invasive niche EGFR^+^ SOX^+^, Type^Inv^) from a hyper-aggressive necrotic niche (ERBB3^+^, Type^Nec^).
- **Vascular Loop and DAMP Cascade:** TypeNec cells deploy a CCL5/CXCL2 chemokine milieu driving perivascular neutrophil recruitment and MPO^+^CitH3^+^ NETosis, sparking a feed-forward loop of hypoxia and necrosis driven by tumor-derived HMGB2^high^ that is pharmacologically targetable.

*Online Key Points (85 characters):*

- Preoperative MRI biomarker of necrotic threshold determines long-term prognosis.
- Cross-species co-mutation profile yielding distinct invasive and necrotic niches.
- **N**eutrophil extracellular traps spark DAMP-targetable feed-forward necroptosis loop.

**Importance of the Study:** Tumor necrosis is a hallmark diagnostic feature of glioblastoma, historically characterized as a passive consequence of local tissue ischemia. This study redefines glioblastoma necrosis as an active, genetically driven immunopathological process. By integrating macroscopic clinical MRI radiomics from over 1,000 patients with single-cell spatial transcriptomics and a novel co-mutant mouse model, we reveal that cell-intrinsic *TP53*, *PTEN*, and *NF1* co-mutations actively dictate a hyper-aggressive necrotic niche. Mechanistically, these mutated tumor cells deploy a CCL5/CXCL2 chemokine cascade to recruit neutrophils, triggering an overactive neutrophil extracellular trap (NET) and a HMGB2-mediated damage-associated molecular pattern (DAMP) loop that accelerates tissue destruction and myeloid immunosuppression. Crucially, we demonstrate that this pathogenic axis can be tracked non-invasively via standard neuroimaging and pharmacologically dismantled with anti-inflammatory small molecules, providing immediate diagnostic stratifications and a brand-new therapeutic vulnerability to disrupt the glioblastoma microenvironment.

**Lay Summary:** Dead tissue zones inside glioblastoma brain tumors, known as necrosis, are tied to poor patient survival. While long believed to be an accidental result of rapid tumor growth cutting off oxygen, this study reveals that tumor cells actively trigger this cell death. Driven by specific genetic mutations, these cells release distress signals that recruit immune cells called neutrophils. These neutrophils create a destructive, inflammatory loop that worsens the disease. By combining patient MRI scans with advanced spatial tissue mapping, we show that this destructive process can be tracked non-invasively and stopped using anti-inflammatory drugs, offering a new treatment strategy.

## Introduction

Human glioblastoma (IDH^wild^ ^type^ ^(wt)^, WHO grade 4) is the most common malignant primary brain tumor in adults. Overall, the prognosis for glioblastoma patients remains poor, with a median survival of less than two years ^1^. Glioblastoma molecular subtypes have been reclassified into mainly classical, mesenchymal, and proneural bulk transcriptional subtypes. However, IDH^wt^ glioblastomas commonly transition from classical to mesenchymal ^2^. Moreover, single-cell studies support that individual tumors may contain a spectrum of transcriptomic and hybrid cell states from multiple subtypes ^3^. The integration and plasticity of these cell states may be linked to cell survival mechanisms, metabolic reprogramming, specific genetic and epigenetic drivers, and the developmental hierarchy of the tumor initiating cell(s) ^4^.

Tumor necrosis is a cell death/survival mechanism associated with chronic inflammation, metastasis, and poor prognosis ^5^. Necrosis is traditionally thought to occur passively when tumor growth outpaces nutrient supply. This notion of irregulated necrosis has been challenged with studies revealing that apoptosis-resistant clones, fueled by intratumor heterogeneity, utilize immunogenic forms of cell death, such as necroptosis, to instruct immune cells in the tumor microenvironment (TME) ^6^. Indeed, neutrophil subsets were shown to actively induce regulated pleomorphic tumor necrosis ^7^. Tumor-associated neutrophils (TANs) are involved in tumor initiation, proliferation, or metastasis, and their presence correlates with poor outcomes ^8^. Uniquely, brain TANs in humans and mice differ significantly from circulating blood neutrophils such as having a prolonged lifespan, immune-suppressive properties, and pro-angiogenic capacity ^9^. Neutrophils exert their functions through chemotaxis, phagocytosis, degranulation, or forming neutrophil extracellular traps (NETs) ^10^. Unlike the passive necrotic cores, neutrophil necroptosis is a programmed, caspase-independent and RIPK3–MLKL-mediated cell death associated with NET-containing chromatin-decondensed DNA aggregates ^10^. Human glioblastoma frequently form a unique configuration necrotic foci, making a dense row of hyperchromatic elongated pseudopalisading cells around a central vascular lumen and/or area of necrosis ^11^. Pseudopalisades are severely hypoxic, overexpress hypoxia-inducible factor (HIF-1) and secrete proangiogenic factors such as VEGF and IL-8 ^11^.

In the adult mouse brain, the reciprocal interactions between tumor initiating cells (TICs) and TME define tumor progression. Neural progenitor cells (NPCs) are continually produced in the subventricular zone (SVZ). NPCs exhibit self-renewal and terminal differentiation, while oligodendrocyte progenitor cells (OPCs) are broadly arrayed in the brain, and generate myelin-forming oligodendrocytes. Both NPCs and OPCs have shown TIC properties. While individual mutations in Trp53, Pten, or Nf1 previously failed to generate penetrant gliomas, compound mutations induce high-grade mouse glioblastomas that faithfully phenocopy the human malignancy ^12–14^. To study glioblastoma cell states and the crosstalk of TICs with the TME, we asked whether ablation of the same set of mouse tumor suppressors; the glioblastoma-prevalent *Trp53*, *Pten*, and *Nf1* conditionally, in adult bipotential progenitors; the NPCs and OPCs, would result in different glioblastoma phenotypes or reproduce glioblastoma necrosis features. To specifically target adult progenitors, we used CreER transgenic mice that drive recombination using the regulatory elements of the mouse *Ascl1* (Achaete-scute family bHLH transcription factor 1; also known as Mash1) in the brain ^15^. Here, we identified NET signatures in large cohorts of glioblastoma patients and generated fully penetrant genetically engineered mouse models (GEMMs) of glioblastoma revealing NET-associated neuroinflammatory signals driving necroptosis, offering new therapeutic vulnerabilities to dismantle the glioblastoma necrotic niche.

## Materials and methods

### Experimental design

The experimental conditions and animal studies were performed in accordance with the institutional animal care and use committee–approved protocols at University of Colorado Anschutz (#01209). Monitoring of the phenotypes of mouse phenotypes, histological analyses, IHC, and histology studies were based on established protocols ^16^, and described in detail in Supplementary Materials.

### Human Subjects and survival analysis

De-identified glioblastoma WHO CNS grade 4 glioblastoma samples were obtained from consented patients at both the UPENN and UCSF biobank and analyzed under a protocol approved by the University of Colorado Anschutz (COMIRB #013-3007). Overall survival (OS) was calculated from the date of preoperative MRI to death or last follow-up among a total of 1,079 glioblastoma patients (585 from the UPENN cohort and 494 from the UCSF cohort). Patients were stratified into “low necrosis” and “high necrosis” groups based on the median percent necrotic core volume (18.57%, n = 1,079), as determined from multi-compartment tumor segmentations (necrotic core, enhancing tumor, and edema) generated from multiparametric preoperative T1 post-contrast and FLAIR MRI sequences ^17^, and described in detail in Supplementary Materials.

### Human Subjects and survival analysis

To evaluate somatic co-alteration patterns in glioblastoma, genomic mutation profiles were assessed within an IDH-wild type cohort (AACR Project GENIE, n=2,850 independent glioblastoma samples). Target gene evaluation focused on key pathways disrupted in canonical glioblastoma: TP53, PTEN, and NF1 (PPN). Pairwise mutational dependency and multi-gene co-occurrence were calculated using a permutation framework modeled after the SelectSim algorithm ^18^, and described in detail in Supplementary Materials.

### Statistical analysis

Data are presented as means ± SEM unless otherwise specified. Seurat FindMarkers with the default Wilcoxon rank-sum test was used for cell marker identification. The Benjamini-Hotchberg procedure was used to make corrections for multiple testing. Survival curves were analyzed using the Kaplan‒Meier survival analyses. A P value of <0.05 was used to denote statistical significance unless specifically indicated. Statistical analyses were performed using Prism version 10.0 (GraphPad).

**See additional Materials and Methods in Supplementary data.**

## Results

### High tumoral necrosis is associated with worse overall survival in glioblastoma patients

To evaluate the prognostic significance of tumor necrosis in diffuse glioblastoma, we analyzed multi-compartment tumor segmentations from preoperative MRI in 1,079 patients from the UPENN and UCSF cohorts. To account for the heterogeneity in necrosis levels, we performed a distribution based optimal percentage cut offs (Supplementary Figure 1). Patients were stratified into high-and low-necrosis groups based on the median percent necrotic core volume (median cutoff = 18.57%), as determined by necrosis density distribution and maximally selected ranked statistics on segmented MRI images (Supplementary Figure 1). Kaplan–Meier survival analysis demonstrated a highly significant separation between the two groups. Kaplan–Meier analysis demonstrated that high tumor necrosis is significantly associated with inferior overall survival despite variabilities among patient cohorts (Supplementary Figure 1). Patients with high tumor necrosis (n = 540) exhibited a significantly shorter overall survival compared with those harboring low tumor necrosis (n = 539) (Log-rank test, p < 0.0001). The survival curves diverged early, but separating beyond the first two years after diagnosis and in long-term follow-up (Figure 1A-C). Like an earlier imaging analysis on a smaller cohort ^19^, glioblastoma patients with extensive necrosis were strongly associated with poor progression-free survival (PFS) and overall survival (OS) (Figure 1C; Supplementary Figure 1B, D, F). High necrotic burden could therefore predict a more aggressive biology and likelihood of diminished responsiveness to therapies. These findings suggest that the extent of tumor necrosis, readily quantifiable on routine preoperative MRI, serves as a powerful non-invasive prognostic biomarker.

**Figure 1.**
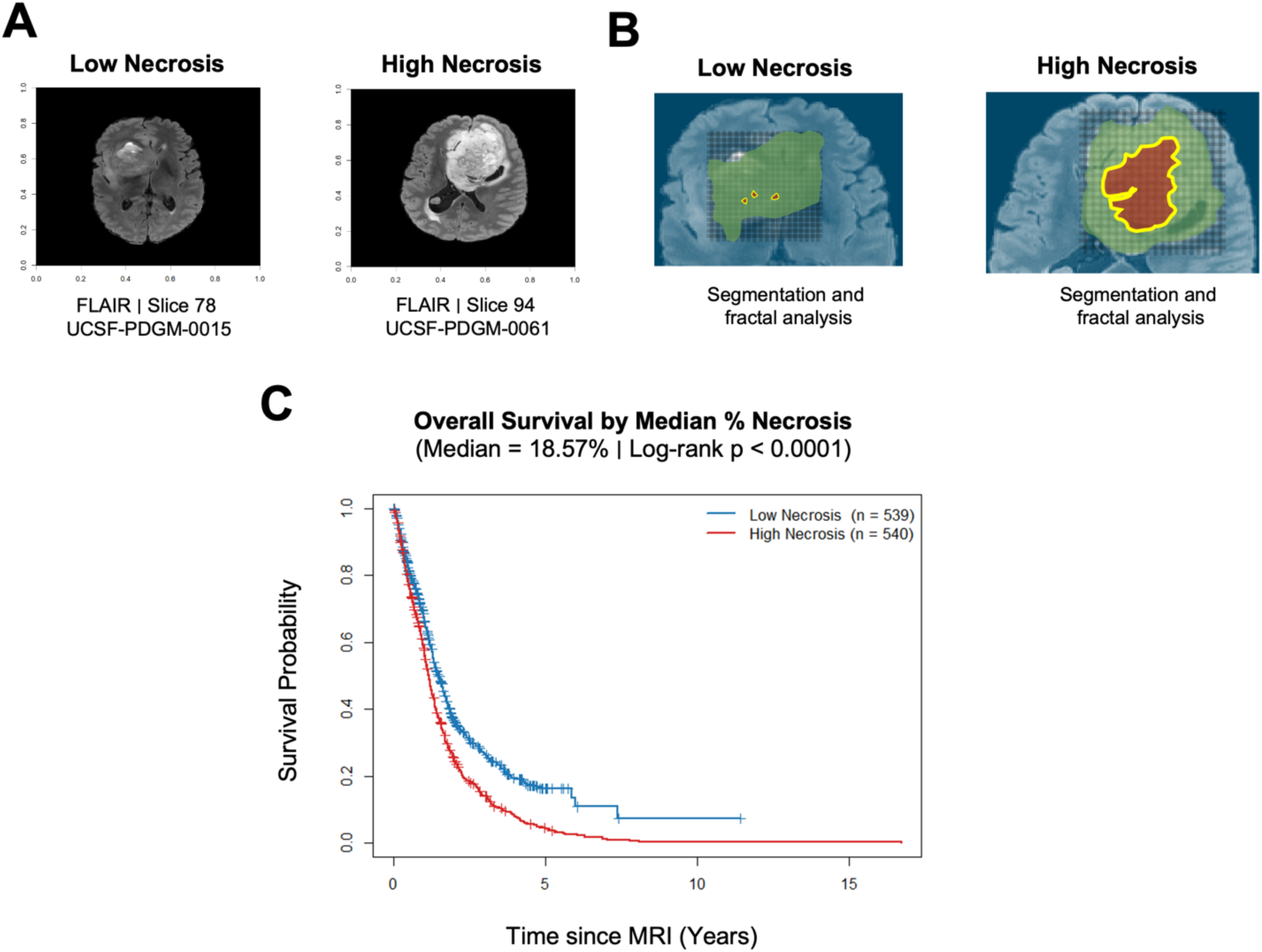
glioblastoma patients with high tumor necrosis have significantly lower overall survival rate. (**A**) Representative post-contrast T1-weighted magnetic resonance image (MRI) from low and high necrosis. (**B**) Segmentation and fractal analysis of the tumor and peritumoral region (T2/FLAIR) (green mask), necrotic ROIs (red mask), and enhancing tumor (yellow). The necrotic ROIs are extracted with the peritumoral ROIs as the border for fractal analysis. Representatives from n = 1,079 MRI images. (**C**) Kaplan–Meier survival curves stratified by median necrotic core percentage (18.57%) in patients with diffuse glioblastoma from the UPENN and UCSF cohort (n = 1,079). Patients with high necrosis (≥18.57%, red, n=540) demonstrated significantly shorter overall survival compared with those with low necrosis (<18.57%, blue, n=539). Log-rank test, P < 0.0001.

To assess if necrosis is tumor cell derived, we next examined our cohort of previously generated mouse brain orthotopic xenografts derived from glioblastoma spheres and patient derived organoids (PDOs) ^20,21^, and identified the originating patient glioblastoma tissues demonstrating either pseudopalisading necrosis or not. We examined the ability of these human glioblastoma cells to propagate the necrosis phenotype in an immunodeficient brain of orthotopic xenografts. Indeed, human glioblastoma cells were either invasive to the surrounding tissue, or propagating a necrosis phenotype, the later only when derived from an original tumor with a pseudopalisading necrosis (Supplementary Figure 2A-C). These initial findings suggest that tumor necrosis could be mechanistically driven from within the tumor cells (Tumor cell-derived necroptosis). To evaluate whether specific somatic mutations may be associated with these phenotypes, and since the original glioblastoma that propagated the necroptosis phenotype displayed mainly a mesenchymal cell state, with notable mutations in TP53, PTEN and NF1, collectively called PPN, we examined the overall co-mutation profiles within IDH-wild type human glioblastoma (AACR Project GENIE, n = 2,850). We first performed multi-gene permutation simulation model, and data suggested that compound mutations of NF1 with TP53 might have an evolutionary co-mutation benefit (Supplementary Figure 2D). Next, we extended the co-mutation analysis to 9,824 patients among multiple cBioPortal cohorts and identified prevalent co-mutations in TP53, PTEN, and NF1 (PPN) in 39%, 34%, and 18%, respectively (Figure 2A; Table S1-S6), and altered PPN were significantly more associated with patients self-identified as White (Figure 2B), with higher mutational counts (Figure 2C), and BRAF, FGFR3, and EGFR genomic changes and/or amplifications, relative to unaltered samples (Figure 2D; Table S2). Strikingly, glioblastoma patients with compound PPN co-mutations exhibited a significantly markedly shorter overall survival compared with the unaltered group (n = 174) (log-rank test, p < 0.0001). These data suggest that in human glioblastoma, PPN somatic co-mutations represent a high-risk group and glioblastoma tumor cells with PPN co-mutations might be functionally relevant in determining invasive or necroptotic phenotypes.

**Figure 2.**
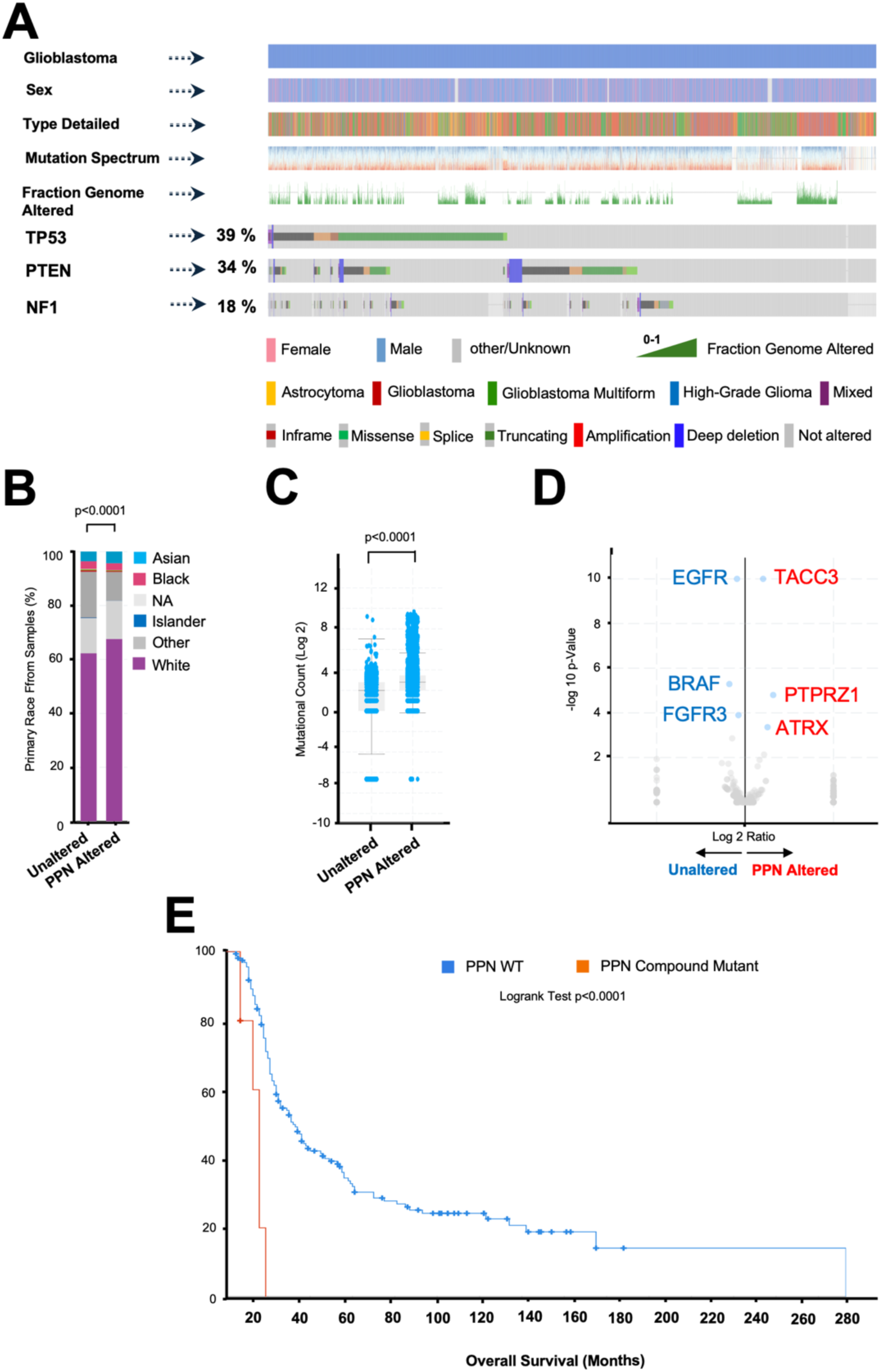
Glioblastoma patients with PPN somatic mutations have significantly lower overall survival rate. (**A**) A visual summary of key clinical features and somatic mutational burden of TP53, PTEN, NF1 (PPN) alteration across glioblastoma patients in the GENIE and cBioPortal databases. An oncoplot analysis of glioblastoma tumor samples (n = 9,824 patients), among these, the percentages of glioblastoma patients that have at least one variant of altered PPN genes are indicated. Original data were queried and downloaded from the cBioPortal for Cancer Genomics. (**B**) Percentages of patient primary race (self-identified and sample recorded). (**C**) Box correlation plot showing mutational count (Log2 score) in PPN mutant and unaltered glioblastomas. Two-tailed unpaired t-tests were used for statistical comparisons. (**D**) Volcano plot showing differentially expressed genomic changes (DEGs) between PPN altered and unaltered, with top structural changes and/or amplifications in *EGFR*, *BRAF*, and *FGFR3* significantly associated with the unaltered group, while PPN alterations were associated with *TACC3*, *PTPRZ1*, and *ATRX* alterations. (**E**) Kaplan–Meier survival curves stratified by compound PPN somatic mutations *vs.* unaltered in diffuse glioblastoma patients (GLASS consortium ^2^) (n = 174). Log-rank test, P < 0.0001.

### GEMM of highly penetrant glioblastoma reveals invasive and necrotic tumor phenotypes

To investigate the molecular mechanisms driving necroptosis in glioblastomas, we utilized a conditional ablation model of the PPN mouse somatic genes. We bred Ascl1-creER mice to incorporate conditional floxed alleles of *Trp53, Pten, Nf1* (PPN) to homozygosity, and fluorescent Brainbow to yield the stable line: Ascl1-creER; Trp53^flox/flox^; Pten^flox/flox^; Nf1^flox/flox^; Brainbow^+/-^ (APPNB). (Supplementary Figure 3A-B). Adult mutants and controls were induced with tamoxifen or vehicle at ∼6 weeks of age. These PPN tumor suppressor deletions were shown to efficiently generate glioblastomas with a complete penetrance ^22^. Since the Ascl1-creER reporter targets two different populations of adult lineage-restricted CNS progenitors, the NSCs and OPCs, these models allowed us to directly assess the functional contribution of Ascl1-lineage cells to glioblastoma initiation and progression (Figure 3A). Mice with individual or dual tumor suppressors did not show phenotypes upon induction and were similar to controls, in accordance with previous reports ^12–14^. APPNB mice showed accelerated morbidity. Conditional compound PPN deletion resulted in a highly penetrant tumorigenesis, with all animals developing neurological symptoms, symptomatic disease and reaching humane endpoints by approximately 4.5 months post-CreER activation (p = 0.0008; Figure 3B). Histological examinations demonstrated expression of Brainbow fluorescent reporters in the white matter and subcortical gray matter regions, including corpus callosum, hypothalamus and thalamic regions, where OPCs and their progeny are located, associated with glioblastoma formation (Figure 3C). While we did not detect any tumors or reporter expression upon histological examination of brain tissues from control mice (Supplementary Figure 3C), H&E staining of APPNB mouse brain sections showed the classical features of diffusely infiltrating high-grade glioblastomas, as determined by neuropathological examination (Figure 3D). Glioblastomas demonstrated ill-defined tumor borders, pseudopalisading necroptosis, and prominent mitoses (Figure 3D). APPNB tumors were classified based on the World Health Organization (WHO) classification system of human CNS malignancies ^23^ as to representing WHO grade 3 astrocytoma or WHO grade 4 glioblastoma.

**Figure 3.**
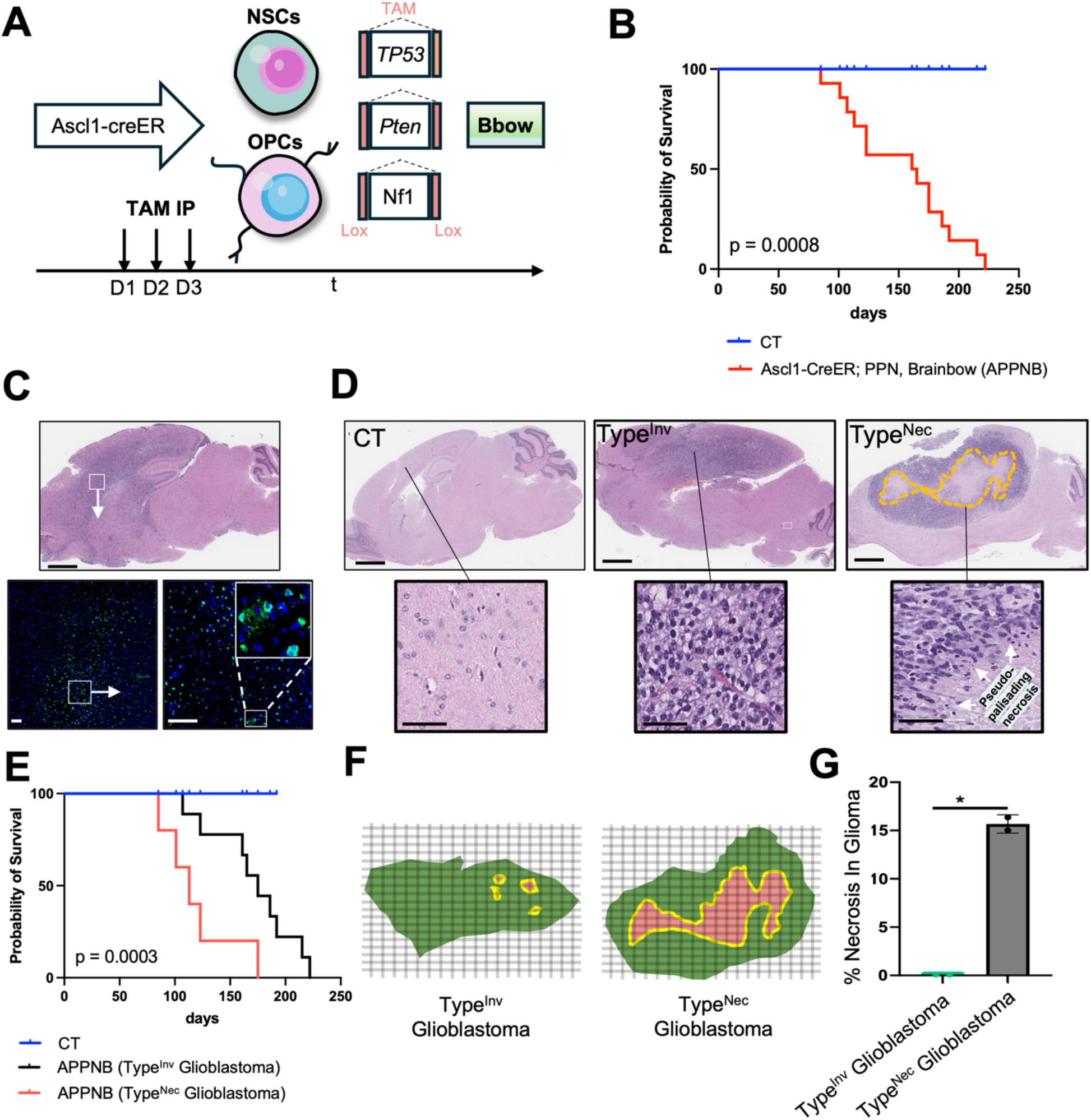
Induction of glioblastomas with PPN deletion using the Ascl1-creER reporter. (**A**) Schematic representation of PPN ablation using Ascl1-creER targeting both NSCs and OPCs. (**B**) Kaplan-Meier survival curves of *Tp53*^-/-^, *Pten*^-/-^, *Nf1*^-/-^, Ascl1-creER^+/+^ mice (n=11) and control mice (n = 10) with recombination induced with tamoxifen IP injection. Survival analysis comparing tamoxifen injected ANPPB mice with control and non-tamoxifen injected (Wilcox log-rank test) (p=0.0008) (**C**) Antibody staining of the fluorescent Brainbow reporters at sites of OPC expression near white matter and subcortical gray matter regions. (**D**) Representative H&E staining images of APPNB brain sections showing a subgroup of glioblastomas with an invasive or necrotic phenotypes. Insert of boxes mark tumor regions with higher magnification, invasive tumor (center) and necrotic tumor (right) relative to normal control (left). (**E**) Kaplan-Meier survival curves of a cohort of mice from the induced APPNB (n=24), with histological diagnosis of stage 4 glioblastomas and either invasive tumors (n=9), or necrotic tumors (n=5) relative to control mice (Wilcox Log-rank test) (p=0.0214). (**F**) Segmentation and fractal analysis of the mouse glioblastoma and peritumoral region (green mask), necrotic ROIs (red mask), and enhancing tumor (yellow). The necrotic ROIs are extracted with the peritumoral ROIs as the border for fractal analysis. (**G**) Percentage of cells in the ROIs with necroptosis from Type^Inv^ and Type^Nec^ glioblastomas (n=10 sections), two-tailed unpaired t-test. Scale bars are 300 μm in D and 60 μm in lower images.

Strikingly, histopathological examination of brain sections revealed two distinct and spatially segregated tumor phenotypes: a highly infiltrative, diffuse glioblastomas predominantly localized to the dorsal brain regions (Type^Inv^) (Figure 3D) and glioblastomas with extensive necrotic areas concentrated in the ventral brain (Type^Nec^) (Figure 3D). Notably, Kaplan–Meier survival analysis further demonstrated a statistically significant difference in the overall survival between the two phenotypes (Figure 3E), with the necrotic phenotype associated with a more aggressive disease progression (Figure 3E-G). Collectively, these results establish the APPNB mouse model as a robust platform for studying the molecular and cellular basis of necroptosis induction in glioblastomas.

### Molecular differences of invasive glioblastomas and necrotic glioblastomas

ASCL1-CreER induces mouse glioblastomas that exhibit marked pathophysiological heterogeneity, appearing as either invasive or necrotic tumors. This heterogeneity is consistent with lineage-dependent glioblastoma subtypes ^22^, demonstrating that mutations in distinct adult CNS progenitors generate molecularly different glioblastomas.

Molecular characterization of the ASCL1-CreER-induced glioblastomas reveals clear subtype-specific marker patterns. ERBB3 (HER3) expression is essentially restricted to the necrotic Type^Nec^ glioblastomas (Supplementary Figure 4A), with being minimal or negligible in Type^Inv^ (Supplementary Figure 4B). In contrast, EGFR (Supplementary Figure 4C-D) and GFAP (Supplementary Figure 4I-J) expressions are significantly upregulated in the Type^Inv^ invasive tumors relative to Type^Nec^. Further supporting this assignment, SOX9 (Supplementary Figure 4E– F) and OLIG2 (Supplementary Figure 4G–H) are expressed at markedly higher levels in the invasive tumors, findings that are independently confirmed by Western blotting (WB) (Supplementary Figure 4K-L). These data further stratified our glioblastoma subtypes into Type^Inv^ (NSC/astrocyte-lineage origin) and Type^Nec^ (OPC origin) based on the established marker profiles

### Proliferative tumor cell phenotypes between invasive and necrotic glioblastomas

To further characterize the phenotypic differences between invasive (Type^Inv^) and necrotic (Type^Nec^) glioblastomas, we evaluated proliferation, sphere-forming capacity, and *in vivo* tumorigenicity. Ki-67 staining demonstrated a significantly higher percentage of proliferating cells in invasive tumor tissues than in necrotic tumors (Supplementary Figure 5A-B). This finding was corroborated by *in vitro* proliferation assays, which showed that tumor cells isolated from invasive glioblastomas exhibited greater proliferative capacity than those isolated from necrotic glioblastomas (Supplementary Figure 5G).

Sphere-forming assays further indicated that invasive glioblastoma cells possessed enhanced self-renewal potential. Tumor cells isolated from necrotic glioblastomas are able to form spheres significantly larger in size than isolated tumor cells invasive glioblastomas (Supplementary Figure 5C-F). However, there is no significant variation in the number of sphere forming ability between isolated invasive and necrotic glioblastoma cells (Supplementary Figure 5F).

Notably, despite their lower proliferation and sphere-forming ability *in vitro*, tumor cells derived from necrotic glioblastomas generated significantly larger subcutaneous tumors upon implantation in mice compared with cells from invasive glioblastomas (Supplementary Figure 5H). These results suggest that while invasive glioblastoma cells display higher proliferative and self-renewal activity under standard culture conditions, necrotic glioblastoma cells possess superior *in vivo* tumorigenic potential.

### Type^Nec^ glioblastomas are characterized by a superior engraftment and distinct secretory profile enriched in pro-tumorigenic and immunosuppressive factors

To further characterize these glioblastoma TME, we isolated tumor cells from both invasive and necrotic glioblastomas. The purity of the isolated populations was rigorously validated by flow cytometry (Figure 4A), leveraging the Brainbow fluorescently-labeled tumor cells. This analysis demonstrated that the isolated invasive and necrotic tumor cells were approximately 99% pure transgene driven-positive populations (Figure 4B). To determine how glioblastoma cells actively shape the immune microenvironment, we isolated cellular protein extracts from purified invasive glioblastoma cells (Type^Inv^) and necrotic glioblastoma cells (Type^Nec^) and analyzed them using a proteome profiler mouse cytokine array (Figure 4C). The assay revealed distinct secretory profiles between the two populations (Figure 4D-E). Notably, extracts from necrotic glioblastoma cells (Type^Inv^) contained substantially higher levels of CXCL2, a key chemokine that mediates neutrophil recruitment. In parallel, several factors critical for recruitment and survival of macrophage, such as MCSF, CCL5, and CXCL12/SDF1 were significantly elevated in necrotic (Type^Nec^) compared with invasive (Type^Inv^) glioblastoma cells. Necrotic cells also showed markedly increased expression of the adhesion molecule CD106 (VCAM1), as well as CCN4 (WISP1), endostatin, and IGFBP3. These factors collectively support neutrophil and macrophage infiltration, endothelial activation, and matrix remodeling within the hypoxic necrotic glioblastoma microenvironment niche.

**Figure 4.**
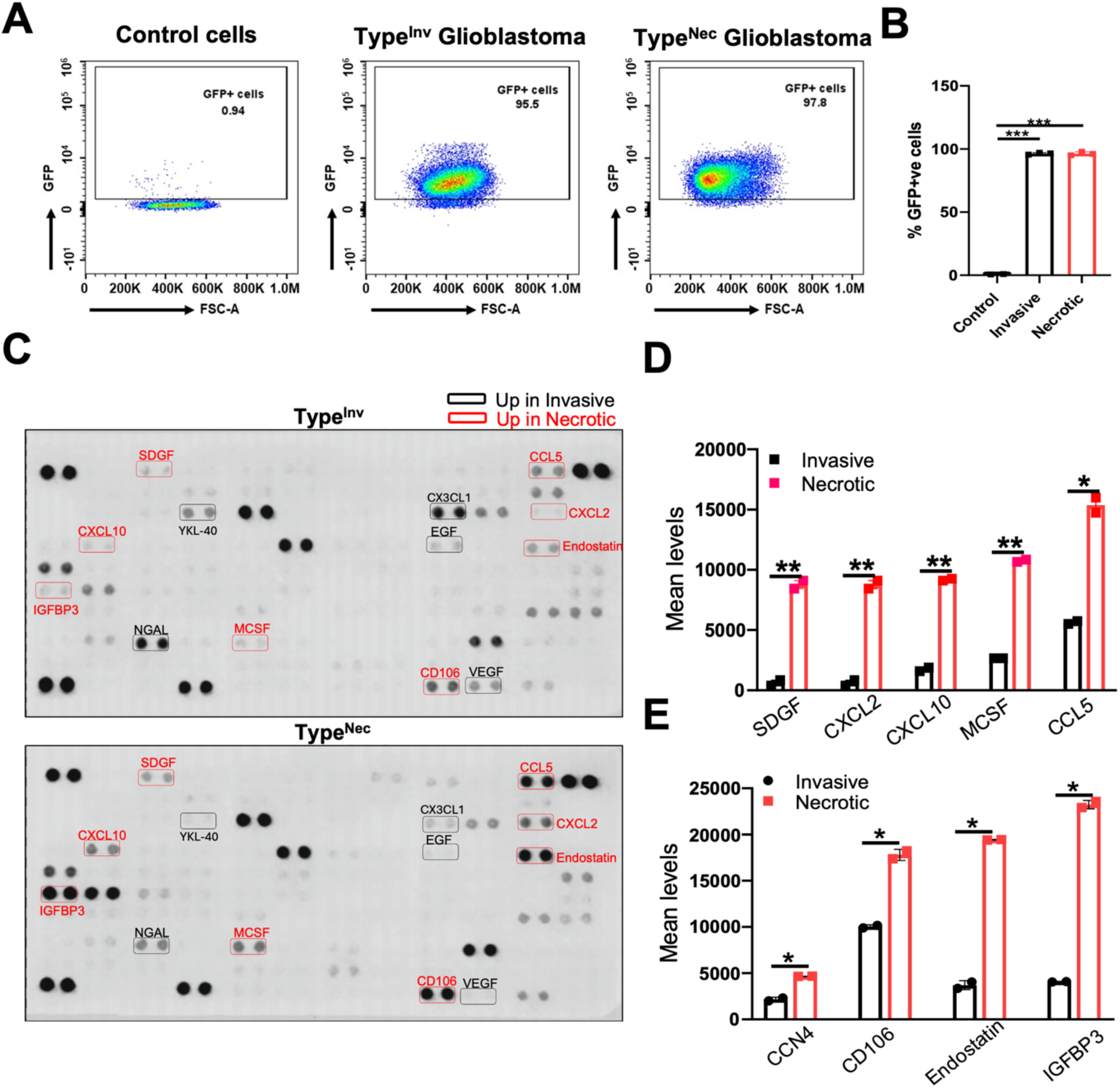
Intra-tumoral cytokine profile of invasive (Type^Inv^) and necrotic (Type^Nec^) tumors. (**A)** The purity of isolated tumor cells from glioblastomas were confirmed by flow cytometric analysis in control (left panel), invasive tumor cells (center panel), and necrotic tumor cells (right panel). **(B)** Representative histogram with percentage of fluorescent cells in the invasive tumor cells (black), and necrotic tumor cells (red). (**C**) Analysis of intra-tumoral cytokine profile between invasive tumor cells and necrotic tumor cells. Immune blot from invasive (top) and necrotic (bottom) glioblastoma cells, with significantly upregulated tumor promoting factors (EGF, VEGF, CGP39, CX3CL1, and NGAL) in invasive tumor cells circulated with black. Red circled cytokines (SDGF, CXCL2 CXCL10, MCSF, CCL5, CCN4, CD106, ENDOSTATIN, and IGFBP3) are significantly upregulated in the necrotic tumor cells. (**D**) Representative histogram of intensity of chemoattractant cytokines (SDGF, CXCL2 CXCL10, MCSF, and CCL5) in invasive (black) and necrotic (red) glioblastoma tumor cells. (**E**) Representative histogram of intensity of tumor promoting cytokines (CCN4, CD106, Endostatin, and IGFBP3) in invasive (black) and necrotic (red) glioblastoma tumor cells. Representative data are shown and expressed as the mean ± SEM, \**p* < 0.05, ** *p* < 0.01, and *** *p* < 0.001.

### Necrotic glioblastomas are enriched with neutrophils that are associated with blood vessels

To investigate the mechanisms underlying the chemokine-enriched microenvironment of necrotic glioblastomas, we focused on CCL5, the most significantly upregulated in necrotic glioblastomas. CCL5 (also known as RANTES) is a pro-inflammatory chemokine within the CXCL2/CXCR2/IL8 axis that attracts immune cells. CXCL2 is also significantly upregulated, when combined; CCL5 and CXCL2 form a multi-layered immunosuppressive shield, with CCL5 populates the tumor with suppressive macrophages, while CXCL2 brings in neutrophils and granulocytic MDSCs ^9^. Immunohistochemistry (IHC) using MPO (a neutrophil marker) and CitH3 (a marker of neutrophil extracellular trap (NET) formation) revealed that relative to invasive glioblastomas (Supplementary Figure 6A–C), a strikingly higher density of MPO⁺ CitH3⁺ neutrophils throughout necrotic glioblastomas (Supplementary Figure 6D-I).

To determine whether these neutrophils physically interact with the vasculature, we performed dual IHC for CD31 (endothelial marker) and MPO. We found CD31^+^ cells associated with microvascular proliferation (MVP) forming glomeruloid vascular structures (Supplementary Figure 7D-F), a definitive histopathological hallmark of glioblastoma. Moreover, necrotic tumors show a significantly higher number of MPO⁺ neutrophils in close proximity to or directly associated with CD31⁺ blood vessels (Supplementary Figure 7A-H). These findings corroborate previous data showing that neutrophil density in the vessel-adjacent zone was higher in glioblastoma tissue than the surrounding normal brain ^9^, and demonstrate that the distinct chemokine profile of necrotic (Type^Nec^) glioblastomas strongly promoting both the recruitment and vascular localization of neutrophils. Consistent with recent studies showing that neutrophils drive vascular occlusion, intratumoral hypoxia, and necroptosis through NET formation, we observed abundant perivascular and intravascular MPO⁺ CitH3⁺ neutrophils observed in necrotic tumors (Figure 4F, 4I), which are likely contributing to microvascular blockage and hypoxia, thereby reinforcing tumor necroptosis in a feed-forward loop. Collectively, our data indicates that CXCL2-and CCL5-mediated neutrophil infiltration and Netosis could be a key mechanism by which necrotic glioblastomas establish and sustain their aggressive, hypoxic, and immunosuppressive microenvironment.

### Macrophage polarization and neutrophil balance are shaped by the TME

CCL5/CCR5 signaling is a key regulator of macrophage recruitment and polarization in the glioblastoma microenvironment ^9^. Thus, we further investigated how CCL5 influenced macrophage populations in invasive and necrotic glioblastomas. IHC for CD163 (anti-inflammatory, M2-like macrophages) and CD32 (pro-inflammatory, M1-like macrophages) revealed clear differences. Invasive glioblastomas were markedly enriched for CD163⁺ macrophages (Supplementary Figure 8A–C), whereas CD32⁺ pro-inflammatory macrophages were significantly more abundant in necrotic glioblastomas (Supplementary Figure 8D–F). To further examine the relationship between macrophages and neutrophils, we performed dual staining. CD163⁺ anti-inflammatory macrophages were significantly more abundant in invasive tumors (Supplementary Figure 9A–C), than in necrotic tumors (Supplementary Figure 9D–F; G– H). In contrast, and consistent with our earlier findings, MPO⁺ neutrophils were significantly enriched in neighborhoods of CD163^+^ macrophages in necrotic glioblastomas. Notably, in invasive tumors, the density of CD163⁺ macrophages were comparable to that of MPO⁺ neutrophils, suggesting a balanced presence of anti-inflammatory macrophages alongside infiltrating neutrophils. These results reveal that invasive glioblastomas are characterized by a predominance of anti-inflammatory CD163⁺ macrophages that coexist in balance with neutrophils ^9^, whereas necrotic glioblastomas might favor pro-inflammatory CD32⁺ macrophages alongside abundant neutrophils.

### Single cell spatial transcriptomics of necrotic and invasive glioblastomas

We next compared the single cell transcriptomic profiles of mouse glioblastomas with different CNS cell populations using references from the mouse brain single cell spatial atlas ^24–26^. We examined H&E-stained sagittal brain sections to guide selection of fields of view (FOVs) for SMI ^27^ on consecutive tissue sections. To ensure a comprehensive representation of the entire section, a median of 20 FOVs were selected based on the presence of tumor cells as determined by a board-certified pathologist. We spatially localized nuclear DNA (DAPI), proteins (antibodies against pan-cytokeratin, CD45, CD298, and B2M) and messenger RNA transcripts across 117 FOVs from 16 mouse brain sections. Similar to our recent single cell spatial profiling of mouse immunocytes ^16^, we applied a morphology-based cell segmentation algorithm to define cell boundaries and removed cells with fewer than 60 total transcripts (Methods). This yielded 59,099 high-confidence single-cell profiles with 81.2 mean features/cell and 139.6 mean count/cell. Unsupervised hierarchical clustering was applied to the single cell spatial transcriptomic profiles of control and glioblastomas brain samples including both Type^Inv^ and Type^Nec^ glioblastomas (Supplementary Figure 10A-D). Clustering was validated using cell type-specific markers compiled from published single cell spatial mouse brain atlas ^24–26^ as a reference (Supplementary Table S7-S8). More than 95% of cells were successfully annotated to a distinct cell type (Figure 5A-D; Supplementary Figure 10; Supplementary Table S7-S8).

**Figure 5.**
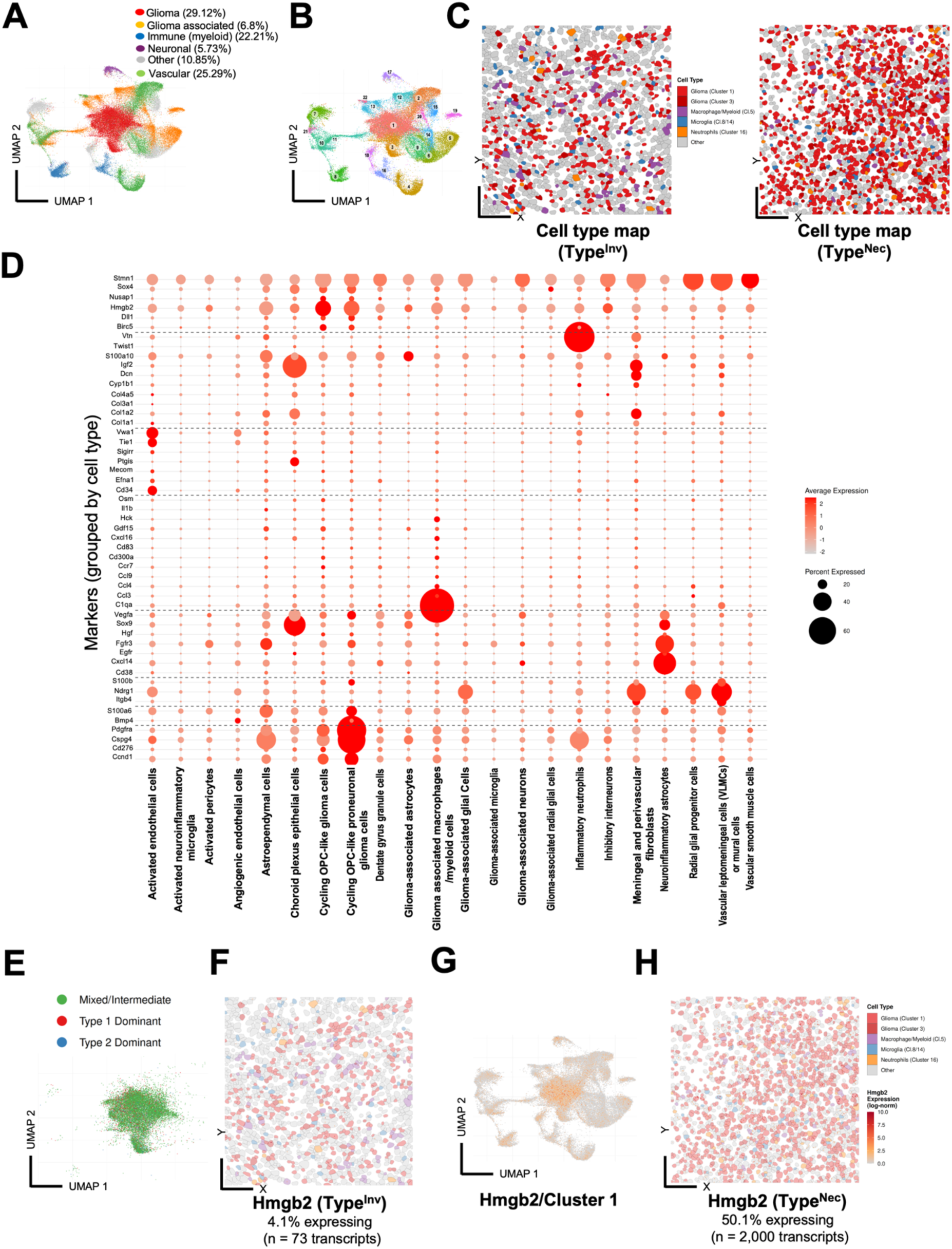
Single cell spatial mapping uncovers tumor and TME cell type diversity in mouse glioblastoma. (**A**) UMAP visualization of mouse glioblastoma tumor cells and TME associated, immune, myeloid, neuronal, vascular, and other cells colored by cell type-annotation across mouse brain sections. (**B**) UMAP visualization of annotated clusters and proportions of the 22 mouse brain cell clusters across samples. Cluster 1 (22.48%), Cluster 2 (9.58%), Cluster 3 (6.64%), Cluster 4 (6.27%), Cluster 5 (5.88%), Cluster 6 (5.49%), Cluster 7 (4.81%), Cluster 8 (4.48%), Cluster 9 (4.45%), Cluster 10 (3.95%), Cluster 11 (3.86%), Cluster 12 (3.52%), Cluster 13 (2.94%), Cluster 14 (2.78%), Cluster 15 (2.46%), Cluster 16 (2.22%), Cluster 17 (2.21%), Cluster 18 (2.03%), Cluster 19 (1.56%), Cluster 20 (1.36%), Cluster 21 (0.69%), and Cluster 22 (0.35%). (**C**) Spatial cell type maps (Cluster 1, light red, cluster 3, dark red), macrophages and myeloid cells (Cluster 5, purple), microglia (Clusters 8 and 14, blue), neutrophils (Cluster 16, orange) and other cells (gray) for representative Type^Inv^ or Type^Nec^ FOVs. (**D**) Bubble heatmap of expression levels of annotated cell types. Color indicates normalized average expression, and dot size indicates the percentage of expressing cells. Cell type markers include Oligodendrocyte/Glioma-associated (*Ccnd1, Cd276, Cspg4, Pdgfra*), OPCs (*Bmp4, S100a6*), Mature Oligodendrocyte (MOL) (*Itgb4, Ndrg1, S100b*), Astrocyte (*Cd38, Cxcl14, Egfr, Fgfr3, Hgf, Sox9, Vegfa*), Microglia (*C1qa, Ccl3, Ccl4, Ccl9, Ccr7, Cd300a, Cd83, Cxcl16, Gdf15, Hck, Il1b, Osm*) Endothelial (*Cd34, Efna1, Mecom, Ptgis, Sigirr, Tie1, Vwa1*), Pericyte (*Col1a1, Col1a2, Col3a1, Col4a5, Cyp1b1, Dcn, Igf2, S100a10, Twist1, Vtn*) and adult NSCs (*Birc5, Dll1, Hmgb2, Nusap1, Sox4, Stmn1*). (**E**) UMAP visualization of glioblastoma cluster 1 with Type^Inv^ dominant (red), Type^Nec^ dominant (blue) or mostly mixed phenotypes (green). (**F**) Spatial cell type map from Type^Inv^ colored by subtype of glioblastoma cells and localized *Hmgb2* RNA transcripts for clusters 1 and 3. (**G**) UMAP visualization of clusters of cells expressing localized *Hmgb2* RNA transcripts (orange). (**H**) Spatial cell type map from Type^Nec^ colored by subtype of glioblastoma cells and localized *Hmgb2* transcripts. Data represent 59,099 cells from 16 mouse brain sections.

Glioblastoma tumor cells (clusters 1 and 3) represented 29.12% of cells, while glioblastoma associated, immune, myeloid, neuronal, vascular and other cell types formed 70.88% of cells (Figure 5A-B). Single cell spatial analysis identified tumor states of OPC-like and proneural (cluster1 and cluster 3, respectively) demonstrating distinct proportions of tumor and TME cell types (Figure 5C), with transitional states (glioma associated glial cells, cluster 11), immune cell contribution to the glioblastoma TME including glioma-associated and neuroinflammatory microglia (cluster 14, 20), M2 TAMs (cluster 8), and activated tumor-infiltrating neuroinflammatory neutrophils (cluster 16), angiogenic endothelial cells (cluster 22), pericytes (cluster 9), and inhibitory and glioma-adjacent stressed neurons (cluster 12, 17) (Figure 5D; Supplementary Figure 10). Single cell spatial maps comparing cell proportions in control and various glioblastomas (Supplementary Figure 11A-B; Supplementary Table S9-S12) demonstrate differentially expressed *Erbb3* transcripts in Type^Nec^ glioblastoma. (Supplementary Figure 11A). Differentially expressed genes (DEGs) from comparison of Type^Inv^ *vs.* Type^Nec^ glioblastomas indicate that Type^Nec^ tumors show higher enrichment in *Hmgb2, Vegfa, Cx3cl1* and *Igfbp3* transcripts relative to Type^Inv^ (Supplementary Figure 12).

To visualize tumor-specific transcript expression among cell types, we performed cell neighborhood matrix analysis of spatial maps for each DEG transcript in glioblastoma clusters 1 and 3 (Supplementary Figure 12; 13) and noted highly DEG in Type^Nec^ *vs.* Type^Inv^, including *Hmgb2* (50.3% *vs.* 4.1% of cells), *Ccl5* (8.2% *vs.* 2.1% of cells), *Vegfa* (10.2% *vs.* 3.1% of cells), *Cxcl3* (12.3% *vs.* 3.8% of cells), *Cxcl2* (4.9% *vs.* 1.8% of cells), and *Cxcl10* (5.6% *vs.* 2.1% of cells) (Supplementary Figure 12; 13), with *Egfr, Sox9, Erbb3, Vegfa, Cxcl1, Csf1, Ccl5, Cxcl2*, and *Hmgb2* largely associated with tumor cell clusters (cluster 1 and 3). These single cell spatial data suggest a necroptosis-associated role for DAMP signaling, predominantly by *Hmgb2*, in the crosstalk between glioblastoma cells and TME in response to pro-inflammatory macrophages alongside abundant neutrophils and NETs to generate the differential immune cell composition, shaped by the distinct chemokine milieu. This mechanistic DAMP signaling activation is validated by demonstrating activation of the pMLKL necroptosis executer in tumor cells associated with NETs and pseudopalisading necrosis (Supplementary Figure 14; 15). To demonstrate that the tumor-cell-derived DAMP signaling could be pharmacologically targeted, we utilized Inflachromene, a microglial inhibitor with anti-inflammatory but without antitumor effects, and Tannic acid, a hERG channel blocker, which suppressed inflammation, and both significantly reduced DAMP *Hmgb1/2* levels (Supplementary Figure 16).

To examine the extent of Type^Inv^ and Type^Nec^ transcriptomic signatures are involved in human glioblastomas, we queried eight human CNS/Brain cancer datasets with profiled mRNA expression levels, including 3,639 glioblastoma patients. We identified the invasive signature of significantly higher EGFR and SOX9 expression at the mRNA (Supplementary Figure 17A-C), protein, and specific EGFR (EGFR^PY1173^ and EGFR^PY1068^ proteins) (Supplementary Figure 17D-E) in the PPN unaltered group, while the necroptosis-associated transcriptomic signature of significantly higher *HMGB2, CCNB1*, and *MPO* are associated with PPN altered glioblastoma (Figure 6A), and significantly more in grade 4 glioblastomas (Figure 6B-C). We stratified glioblastoma samples based on *ERBB3*^high^ *vs. ERBB3*^low^ expression (Supplementary Figure 18A; Supplementary Table S13), and higher *ERBB3* expression is associated with higher DAMP, TNF, myeloid, and inflammatory secretory transcriptome (Supplementary Figure 18B; Supplementary Table S13).

**Figure 6.**
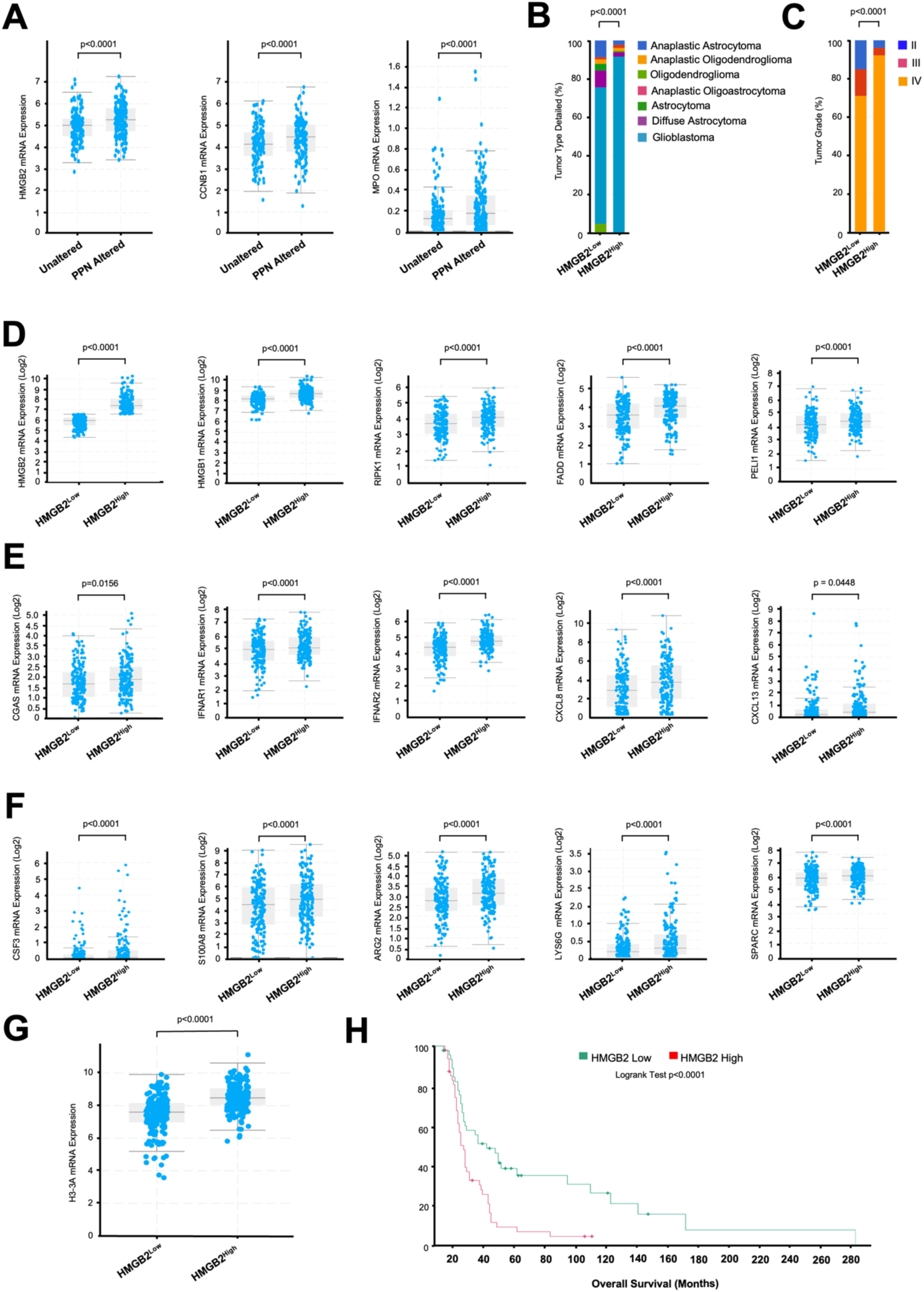
**Necroptosis signature in human glioblastoma patients**. (**A**) Box correlation plot showing *HMGB2* mRNA expression among the GDC dataset (n=523 glioblastoma patients), and significantly higher correlation with *CCNB1* tumor growth signal and *MPO* myeloid signature in glioblastoma patients with PPN altered (n=237 patients) *vs.* PPN unaltered (n=286 patients). **(B)** Percentages of detailed glioblastoma subtypes associated with *HMGB2*^high^ (n=523 patients). Note the near complete absence of oligodendrogliomas among the *HMGB2*^high^ group. Tumor types were derived from original samples and classified according to the 2016 World Health Organization classification of tumors of the central nervous system. (**C)** Bar graphs on the percentages of glioma tumor grades 2-4 among *HMGB2*^high^ and *HMGB2*^low^ groups within the GLASS consortium (n=329 patients). (**D-E**) Box correlation plot showing mRNA expression (Log2 score) among glioblastoma patients within the GLASS consortium ^2^ (n=329 patients) for *HMGB2* with DAMP activation (*HMGB1, RIPK1, FADD*, and *PELI1*), interferon and inflammatory recruitment (**E**). (**F**) Correlation plot of mRNA (Log2 score) signature of neutrophil activation (*CSF3* transcript of GCSF) and myeloid-derived activation of PMN-MDSC (*S100A8, ARG2*, and myeloid *LYS6G*, and *SPARC*). (**G**) Box correlation plot of histone *H3-3A* mRNA expression in *HMGB2*^Low^ *vs. HMGB2*^high^ groups. Two-tailed unpaired t-test were used for statistical comparisons of mRNA correlates. (**H**) Kaplan-Meier survival curve of glioblastoma patients with *HMGB2*^Low^ *vs. HMGB2*^high^ expression (Log-rank Test p<0.001).

To probe the extent of *HMGB2* is tightly linked to the hyper-aggressive, necroptosis-activating state of glioblastoma tumor cells, we examined the correlation between *HMGB2*^high^ and *HMGB2*^Low^ transcriptome with the core factors that are associated with MES-cell state (*CHI3L1* and *CD44*) (Supplementary Figure 19A), not associated with pyroptosis (Less *NLRP3* and *CASP8*) (Supplementary Figure 19B), but significantly higher transcripts associated with DAMP signaling and necroptosis (*HMGB1*, *RIPK1, FADD*, and *PELI1*) (Figure 6D), *C-GAS*-STING, Interferon, TNF, TLR, hypoxia, and inflammatory recruitment signals (Figure 6E; Supplementary Figure 19C-D), myeloid and neutrophil activation and maintenance of glioblastoma immunosuppressive states (*CSF3* transcript of GCSF, *S100A8, ARG2*, myeloid *LYS6G*, and *SPARC* transcripts of activated PMN-MDSCs) (Figure 6F; Supplementary Figure 19C-D; Supplementary Table S14). Moreover, the *HMGB2*^high^ group has significantly higher *H3-3A* (Figure 6G), a biomarker of overactive NETs, prior to citrullination (turning post-translationally into CitH3), associated with glioblastoma tumor aggressive signals (Supplementary Figure 19E). Critically, the overall survival of *HMGB2*^high^ patients was significantly worse (Log-rank test p<0.001) (Figure 6H). These data suggest that the PPN-associated hyper-aggressive, necroptosis-activating state of glioblastoma possesses myeloid derived inflammatory tumor suppressive microenvironment.

## Discussion

Human glioblastomas frequently form a unique configuration of pseudopalisading necrotic foci around a central vascular lumen. We demonstrate that the pseudopalisading necrosis phenotypes are associated with neutrophil necroptosis and show the prognostic significance of necroptosis in large cohorts of patients. The distinction between necroptosis and an invasive edge in glioblastoma is a fundamental pathological diagnostic requirement ^28^. We utilized murine and xenograft models and human glioblastoma to study the complex spatiotemporal events of TME involved in the development of necroptosis. Our APPNB mice reliably mimic human glioblastoma features, including microvascular MVP (glomeruloid structures), highly infiltrative margins, and pseudopalisading necrosis. The proposed mechanisms by which pseudopalisading necrosis occurs were: 1) intravascular thrombosis, 2) hypoxic regions around vascular pathology, 3) outward cell migration creating a peripheral moving wave (pseudopalisade), 4) death of non-migrated cells, 5) secretion of proangiogenic factors by hypoxic cells, 6) microvascular proliferation, and 7) expansion toward new vasculatures ^11,29^. We describe neutrophil necroptosis as a key tumor cell derived mechanism to sustain the dramatic change in glioblastoma cell states. Studies of *IDH*^wt^ glioblastoma demonstrated that differentiated-like cell state positively associated with pseudopalisading cells around necrosis ^2^. In TNBC models, neutrophil subsets were recently shown to actively induce vascular occlusion and cause regulated pleomorphic tumor necroptosis ^30^. We demonstrate that glioblastoma cells, according to glioblastoma cell of origin, modulate the TME through neutrophils, NETs, and distinct chemokine milieu to develop the pseudopalisading necroptotic phenotype.

Although in glioblastoma, the cell of origin remains unresolved, the putative TICs are the OPCs or possibly differentiated neurons. The NPCs and OPCs restricted phenotypes, we targeted with the conditional ablation of PPN, resulted in invasive or necroptotic glioblastoma subtypes. While supported by previous studies ^22^, various TICs might underlie a diversity of functional subtypes found in human glioblastoma. Thus, glioblastoma cell state plasticity could enable malignant cells to not only assume upstream stem-like states but also differentiate into states across the NPCs and OPCs developmental zones. Since more differentiated cell types could be induced to give rise to glioblastoma in mouse models ^31^, and a similarly invasive glioblastoma phenotypes shown to be driven by OPCs though neuron-tumor interactions ^32^, the reciprocal interactions between TIC and TME cells could contribute to heterogeneous cell states ^4^. We identified GVP clusters in our mouse glioblastomas, which are phenotypic hallmarks of the mesenchymal (MES-like) cell states, where *NF1* and *PTEN* deletions are enriched ^3^. Tumorigenic glioblastoma stem cells (GSCs) were shown to give rise to pericytes to support vessel function and tumor growth ^33^, and GVPs may be derived from divergent vascular immune landscapes ^34^, therefore contributing to mass necroptosis by creating the GVP clusters associated with the classic pseudopalisading cores.

Since our APPNB immunocompetent glioblastoma model reliably generates both invasive and necrotic phenotypes within the same genetic background, it is well-suited for identifying driver modulators of the glioblastoma TME. Tumor cells use combined physical barriers, metabolic traps, and chemical reprogramming to control nearby immune cells. Because the PPN co-mutation profile is associated with a predominant MES-like state, the TME becomes highly active, aggressively suppressing immune cells ^35^. Loss of *TP53* removes key transcriptional control over DNA damage responses and apoptosis. Loss of *PTEN* leads to uncontrolled survival signals that cross-talk with inflammation and necroptosis. Additionally, loss of *NF1* elevates RAS, continuously forcing downstream RAF-MEK-ERK cascade. This cascade likely activates *FOSL1*, the master transcription factor genetic driver of MES-like state ^36^. The chronic oncogenic cascade likely shift cells toward forming the necrosome complex. Feed-Forward cross-talk with neutrophils drives the tumor toward greater release of CXCL1/2, as our single cell spatial data shows, pulling neutrophils into the hypoxic core. The neutrophils undergo NET and release TNF-α, reinforcing more MES-like state with a proneural-to-mesenchymal Transition (PMT) ^37^.

The quantity and composition of the TME myeloid cells in glioblastoma are dictated by the genetic driver mutations ^38^. Glioblastoma TME is dominated by immunosuppressive TAMs. We demonstrate that pro-inflammatory macrophages alongside abundant TANs and NETs generate differential immune cell composition, shaped by Type^Inv^ or Type^Nec^ distinct chemokine milieu. Our studies show that DAMP signaling ^29^, predominantly by HMGB2, is linked to TAN-and NET-associated necroptosis. HMGB1, which is widely expressed ^39^, mainly functions extracellularly, while the spatially restricted HMGB2 primarily activate oncogenic signaling and senescence associated inflammation ^40^. We recently identified HMGB2 among pro-tumorigenic senescence-associated secretory phenotype (SASP) signatures of chronic inflammation and tumor formation ^16^. HMGB2 represents an early sign of progression in glioblastoma ^41^, as it promotes T-cell exhaustion ^42^. Our data suggest that chronic DAMP and Hmgb2 activate the necroptosis executer pMLKL and are associated with granulocytic myeloid-derived suppressor cells (G-MDSCs), expressing arginase-1/2 (ARG1/2) with macroscopic NET vehicles that disseminate DAMP signaling. Prior studies suggested that CXCL1-CXCR2 tumor myeloid axis drive necrosis ^30^. NETs stimulate the NF-κB signaling, thus promoting IL-8 secretion, and IL-8 recruits neutrophils for NET formation ^43^. Consistently, patients with elevated circulating MDSCs tend to have elevated serum IL-8 levels ^44^.

We identify a cascade of tumor-ERBB3, and DAMPs interrelate with TANs and NET-induced necroptosis in glioblastoma. Our findings have direct clinical significance. Elevated tumor necroptosis reflect a more aggressive tumor biology, and can serve as a simple, inexpensive pathologic biomarker for early risk stratification. Patients with NETs may benefit from intensified multimodal therapy, closer surveillance, and/or enrollment in trials targeting necrotic pathways. Incorporating quantitative AI and digital necroptosis assessment into prognostic models and pathology reports could improve personalized treatment decisions. Moreover, establishing companion diagnostic tests for circulating NET biomarkers (e.g., serum and/or CSF MPO-DNA or H3Cit-DNA, tumor-educated platelets, or circulating reprogrammed neutrophil) are essential for identifying patients with NET-induced necroptosis and high NET burden in clinical trials.

Single cell proteomics identified ∼1,100 proteins from single TANs of glioblastoma ^45^, offering new therapeutic targets. Therapeutic strategies for targeting NET-induced necroptosis would ideally yield durable responses against glioblastoma cell states ^46^, and include inhibiting hypoxia while sensitizing glioblastoma to TMZ and/or antiangiogenic therapy ^47^, anti-HER3 (ERBB3) antibody to block HER3/NRG1 signaling ^48^, targeting DAMPs/HMGB2, which may not only attenuates tumor growth but also reverses T-cell exhaustion ^49^, and at least four current NET-directed interventions, including 1) inhibiting NET formation with PAD4 inhibitors, 2) increasing NET degradation/clearance using DNase I, 3) blocking CCDC25, MMP9P, TLR9, or siglec-sialic acid axis linking altered tumor glycans to NET-mediated immunosuppression, and 4) NET blockade for priming in combination with immunotherapy ^50^. Critically, in the context of immunotherapy, necroptosis release DAMPs that reshape the immune TME; therefore, identifying patients with high necroptosis provide distinct immunologic features to select those who would respond to immune or myeloid checkpoint inhibitors or combination regimens.

## Ethics

Animal studies were reviewed and approved by the University of Colorado IACUC. For investigations of glioblastoma patients transcriptomics, data were accessed from public repositories, and informed consent was obtained from the participants of the original studies cited.

## Data Availability

MRI imaging [Link 1: https://github.com/data-nih/tcia/blob/main/upenn-gbm.md] [Link 2: https://www.cancerimagingarchive.net/collection/ucsf-pdgm/]
Genomic and clinical glioblastoma data [Link: https://genie.cbioportal.org/].

## Acknowledgments

We thank CU Anschutz Breeding Core for assistance with mouse breeding. We thank UCCC Pathology Shared Resource (PSR) and Genomics Shared Resource (GSR) for tissue arrays and single cell spatial assays.

## Author contributions

Conceptualization: PV, HES

Investigation: PV, LL, HES

Visualization: PV, LL, SI, HES

Funding acquisition: HES

Project administration: HES

Writing – original draft: PV, HES

Writing – review & editing: PV, LL, YZ, SI, SL, BG, AVB, BTH, SRP, HES

## Data availability

All data are available in the main text or the Supplementary information. Computational pipelines and custom R/Python scripts used for spatial transcriptomic alignment will be available for the purposes of reproducing or extending the analysis at: https://github.com/orgs/SabaawyLab/repositories

## List of Supplementary Materials

Materials and Methods

Data Resources

Fig. S1 to S19

Data file Tables S1 to S14 (Excel files)

