## Supplemental data for "Tumor Associated Neutrophils Extend NETs To Spatially Drive Necroptosis In Glioblastoma"

For

**List of Supplementary Materials**

Materials and Methods

Figure S1 to S19

Data file Tables S1 to S14 (Excel files)

### SUPPLEMENTARY MATERIALS AND METHODS

#### Animal models

Animal models and experimental conditions were performed in accordance with the institutional animal care and use committee–approved protocols at University of Colorado Anschutz (#01209). Monitoring of the phenotypes of mouse phenotypes, histological analyses, IHC, and histology studies were based on established protocols [1]. To select mouse models, we considered that the cell of origin of glioblastoma remains unresolved [2], and the putative TICs are the OPCs [3] or possibly differentiated neurons [4]. We utilized CreER transgenic mice that drive recombination using the regulatory elements of the mouse *Ascl1* (Achaete-scute family bHLH transcription factor 1; also known as Mash1) and *GFAP* to target adult brains bipotential progenitors; the NPCs and OPCs using *Ascl1* and astrocytes using *GFAP*, respectively [3,5-7]. All mouse models were maintained on a mixed 129Svj/C57Bl6/B6CBA background. Tumor suppressor floxed mice containing the Ascl1-creER, with tamoxifen (Cat# T5648, Sigma-Aldrich, St. Louis, MO, USA) or vehicle (9:1, sunflower oil: ethanol mixture) by intraperitoneal injection using a working concentration of 27 mg/ mL. Six week-old APPNB mice (Ascl1-creER; *Trp53*$^{flox/flox}$ ; *Pten*$^{flox/flox}$ ; *Nf1*$^{flox/flox}$; Brainbow$^{+/-}$ were administered with 225 mg/kg tamoxifen once a day as a single dose or for 3 consecutive days. Cohorts of *Trp53*$^{flox/flox}$ , *Pten*$^{flox/flox}$, and *Nf1*$^{flox/flox}$ mice were used in parallel to uninjected controls. Mice positive for mice GFAP-creER were crossed with the PPN mutant mice and used in the single cell spatial studies to confirm astrocytic lineages. Mice aged 4-8 weeks were injected intraperitoneally for 3 consecutive days at a dose of 9 mg tamoxifen per 40 g body weight. Overall survival rates were determined by monitoring mice with the desired genotypes twice per week for reaching experimental endpoints, defined by a change in the behavior and decrease in body weight and posture changes.

**Image analyses of MRI from Human Subjects and survival analysis**

De-identified glioblastoma WHO CNS grade 4 samples were obtained from consented patients at both the UPENN and UCSF biobank and analyzed under a protocol approved by the University of Colorado Anschutz (COMIRB #013-3007). Overall survival (OS) was calculated from the date of preoperative MRI to death or last follow-up among a total of 1,079 glioblastoma patients (585 from the UPENN cohort [8] and 494 from the UCSF cohort [9]). Patients were stratified into "low necrosis" and "high necrosis" groups based on the median percent necrotic core volume (18.57%, n = 1,079), as determined from multi-compartment tumor segmentations (necrotic core, enhancing tumor, and edema) generated from multiparametric preoperative T1 post-contrast and FLAIR MRI sequences [10]. The dataset include neuropathologist expert-corrected segmentations of tumor subregions. Tumor regions of interest (ROIs) were drawn by delineating the abnormal hyperintense signals on post-contrast T1-weighted images. Necrotic ROIs were defined as regions with decreased signal intensity within the tumor. Necrotic ROIs of the tumor were extracted with the tumor ROIs at the same slice as the border. Following binarization, the two-dimensional data pertaining to the necrosis slices were loaded in ImageJ software. For fractal analysis, we used the box-counting method with Fraclac. Necrotic core volume was quantified as a percentage of total tumor volume using validated semi-automated segmentation pipelines. Kaplan–Meier survival curves were generated for the low-necrosis (median <18.57%, n = 539) and high-necrosis (≥18.57%, n = 540) groups. Differences in survival distributions were assessed using the log-rank (Mantel–Cox) test. All statistical analyses were performed in R (version 4.3.3) using the survival and survminer packages. Two-sided p-values < 0.05 were considered statistically significant. No patients were excluded from the UCSF dataset survival analysis except for 7 patients with missing necrosis volume data.

**Somatic co-mutation analyses of Human Subjects and survival analysis**

To evaluate somatic co-alteration patterns in glioblastoma, genomic mutation profiles were assessed within an IDH-wild type (IDH$^{wt}$) cohort (AACR Project GENIE, n=2,850 independent glioblastoma samples). Target gene evaluation focused on key pathways disrupted in canonical glioblastoma: *TP53, PTEN,* and *NF1* (PPN). Pairwise mutational dependency and multi-gene co-occurrence were calculated using a permutation framework modeled after the SelectSim algorithm [11]. For each target gene pair (*TP53/PTEN, TP53/NF1*, and *PTEN/NF1*), the true observed simultaneous dual or three-gene co-occurrence mutation frequency was determined. A null distribution was generated via background Monte Carlo simulations (2,000 permutations). During each permutation, individual sample mutational vectors were randomly redistributed while maintaining the fixed marginal mutation rates across the entire cohort. Expected co-mutation frequencies were defined as the mean overlap across all simulated iterations. Empirical p-values (Score P val) were calculated as the proportion of permutations where the simulated co-occurrence met or exceeded the true observed count. Simultaneous concurrent alterations targeting all three pathways (*TP53, PTEN, NF1*) were observed in 1.4% (n = 40) of the glioblastoma cohort. demonstrating that simultaneous coexistence of *TP53, PTEN*, and *NF1* alterations does not deviate significantly from random neutral expectations in this cohort. We next queried eight human CNS/Brain cancer datasets with profiled mRNA expression levels to examine the extent of Type$^{Inv}$ and Type$^{Nec}$ transcriptomic signatures in a large number of human glioblastomas. For our analysis, we retained samples categorized as primary. Next, we excluded samples lacking mRNA Z-scores. Upon these filtering steps, we retained PPN mutation data. Tumor type and subtype annotations for the cBioPortal datasets were collected from publications. Tumor samples were stratified further in mRNA expression levels (no alteration and within normal range mRNA Z-scores), Low expression (Z-score <=-0.4), including missense mutations, which are nucleotide changes that lead to single amino acid substitutions, and deep deletions, which include truncating mutations encompassing several types of putative loss of function mutations such as in-frame and out-of-frame insertions and deletions, nonsense and nonstop mutations, variants in splice site, splice region and translation start site. Since truncating mutations are generally considered loss of function variants, we did not apply further filters on these variants and retained all truncating mutations. We used the Onco Query Language (OQL) [12] to identify samples with alterations in *TP53*, *PTEN*, and *NF1* (PPN).

To determine the expression level cutoff of *ERBB3* and *HMGB2* in human datasets, we reasoned that heterozygous loss (50% dosage) might yield a phenotype due to genetic buffering [13], therefore we established levels above unaltered expression levels (Z-score >=+1.2) as high, and (Z-score <=-0.4) as low. These cutoffs were validated among four additional datasets, by confirming the linear step-down in mRNA expression using a linear trend test, demonstrating no overlaps between the groups, and loss of function variants always associated with samples designated as low expression.

**Sample embedding, immunofluorescence, and Immunohistochemistry**

Organoid and sphere cells were first removed from wells, spun down (5 minutes, 200g, 4°C), and washed in 1mL PBS. Cells were fixed by resuspension in 1 mL 4% PFA/PBS for 45 minutes at 4°C with intermittent mixing. After washing with PBS and centrifugation, cells were embedded in pre-heated Histogel, allowed to solidify, and transferred to a histology cassette in 70% ethanol before sectioning. Formalin-fixed, paraffin-embedded (FFPE) glioblastoma tumor tissues were cut into 4 µm sections. De-paraffinized the slides using xylene and followed by ethyl alcohol (Thermo Fisher Scientific, Rockford, IL, USA) rehydration. Antigen retrieval was performed by placing the sections in citrate buffer (pH 6.0) (Vector Laboratories, Newark, CA,

USA) for 20 min at 120 °C followed by 30 min at room temperature. Tumor sections were blocked with M.O.M blocking reagent and the Vectastain Elite ABC-HRP kit (Vector Laboratories, Newark, CA, USA), and incubated overnight at 4 °C with primary antibodies M.O.M diluent reagent: anti-OLIG2 (1:100; Abcam, Cat: ab1667), anti-Ki-67 (2C5) (1:100; Abcam, Cat: ab1667), anti-cleaved caspase3 (1:200; CST Cat:9661s), anti-ERRB3 (1:200; CST, cat: 12708s), anti-GFAP (1:100; Abcam, Cat: ab68428), anti-SOX9 (1:400; Cat: AB5535), and anti-EGFR (1:100; Abcam, Cat: ab52894). The slides were then stained by Alexa Fluor® 488 labeled secondary antibodies (Invitrogen, Waltham, MA, USA) for 1 h at room temperature, mounted with DAPI Fluoromount-G® (Southern Biotech, Cat: 0100-20, Birmingham, AL, USA) and analyzed using Olympus Fluorescent Microscope IX83 (Leica, Boston, MA, USA).

**RNA Extraction and Quantitative PCR Analysis**

RNA was extracted by QIAGEN RNeasy kit (QIAGEN, Cat: 74136, Germantown, MD, USA) with the manufacturer's protocol. RNA amount was quantified by an ND-1000 spectrophotometer (Nanodrop Technologies, Wilmington, DE, USA). cDNAs were reverse transcribed with total RNA (500–1000 ng per sample) using Reverse Transcription Kit iScript cDNA synthesis kit (Bio-Rad, Cat: 1708891, Hercules, CA, USA). Quantitative PCR was performed using PowerUP SYBR Green PCR Master Mix (Applied Biosystem, Cat: A25742, Waltham, MA, USA), and the samples were amplified with AriaMx Real-time PCR System (Agilent Technologies, Santa Clara, CA, USA). GAPDH were used as in housekeeping control. The qPCR conditions were 50 °C for 2 min, 95 °C for 15 min, followed by 40 cycles of 95 °C for 15 s, 60 °C for 30 s. Primer sequences are available upon request.

**Tumor cytokine expression profile**

The cytokine profiles of the tumor homogenates were analyzed using the Proteome Profiler Mouse XL Cytokine Array Kit (ARY028; R&D Systems, USA) in accordance with the manufacturer's instructions. Briefly, the membranes were blocked with blocking buffer for 1 h, incubated with each sample (200 µg) overnight at 4 °C, and then preincubated for 1 h with a biotinylated detection antibody cocktail. The membranes were subsequently incubated with streptavidin-HRP for 30 min and washed with wash buffer. The levels of cytokines/chemokines were measured by determining the spot intensity using ImageJ software.

**Spheres, organoids and mouse primary cell cultures**

Mouse primary glioblastoma cancer cell lines from glioblastomas developed in Ascl1-cre-ER, $Nf1^{fl/fl}$; $Pten^{fl/fl}$ $p53^{fl/fl}$; $Brainbow^{+/-}$ mice, using mechanical dissociation techniques and cultured in neural basal media (Thermo Scientific, Rockford, IL, USA) supplemented with GlutaMAX (Thermo Scientific, Rockford, IL, USA), heparin (2ug / mL), mouse EGF (20ng/ mL), mouse FGF (20 ng / mL), B-27, N-2 with 100 IU/ml penicillin-100 µg/ mL streptomycin (Thermo Scientific, Rockford, IL, USA) at 37°C and 5% CO2 in the incubator. Each cell line was cryopreserved at early passage numbers (less than six passages after acquisition) and utilized in experiments for up to 20 passages. The ability of these cells to make sphere and organoid cultures were examined as previously described [14]. Organoid models included 10% ECM (Cultrex) [14] and were passaged via macro-dissection into ~1 mm3 pieces according to visual assessment. When required, sphere and organoid models were dissociated in Accutase (STEMCELL Technologies #07920) for 5 minutes at 37°C. Single cells were plated on ultralow-attachment, 6-well plates in cultured in neural basal media (Thermo Scientific, Rockford, IL, USA) supplemented with GlutaMAX (Gemini Bioproducts, West Sacramento, CA, USA), heparin (2ug / mL), mouse EGF (20ng/ mL), mouse FGF (20 ng / mL), B-27 (50X) (Gibco, ), N-2 (100X) (Gibco,) with 100 IU/ml penicillin-100 µg/ mL streptomycin (Mediatech, Manassas, VA, USA) at 37°C and 5% CO2 in the incubator for tumor sphere formation as previously described [15]. After 7 to 14 days of culture, to measure the size of the tumor spheres, images were taken at 5–10 randomly chosen areas under a phase contrast microscope and analyzed by ImageJ software.

**Western blotting**

Cells or tissue were lysed by RIPA buffer (Boston Bioproducts, Inc., Cat: BP-115, Milford, MA, USA) added with protease/ phosphatase inhibitors cocktail (Sigma-Aldrich, Cat: P8340A, St. Louis, MO, USA). Samples were centrifuged at 10,000× g for 10 min at 4 °C, and the supernatant collected. The protein concentration was determined with a DCTM Protein assay (Bio-Rad, Cat: 5000114, Hercules, CA, USA). The lysates were mixed with SDS samples buffer and heated at 95 °C for 5 min. Equal quantities of protein were loaded with SDS-PAGE gels (cat# , Bio-Rad, Boulder, CO) and transferred onto polyvinylidene difluoride membranes (0.4 µm) (Sigma-Aldrich, St. Louis, MO, USA) at 100V for 1 h. The blots were blocked with 2% BSA and incubated with primary antibody for 1h, followed by the secondary antibody for 1 h. Blots were developed with horseradish peroxidase substrate SuperSignal™ West Femto (Thermo Fisher Scientific, Cat: 34095, Waltham, MA, USA) and analyzed by Li-Cor Odyssey M imaging system (LI-COR, Lincoln, NE, USA). Samples for the same experiment were run simultaneously and probed for multiple proteins. Anti-PTEN (Cat: 7649) (1:1000), anti-p53 (Cat: 2524S) (1:1000) and anti-NF1 (Cat: 14623s) (1:1000), anti-SOX9 (Cat: AB5535) (1:1000), anti-EGFR (Cat: ab68428) (1:1000) antibodies were purchased from Cell Signaling (Danvers, MA, USA). Anti-ERBB3 antibody was purchased from Novus Biologicals (Cat: NBP1-76769, Centennial, CO, USA) (1:1000). Anti-Gapdh (Cat: PA1-987, Waltham, MA, USA) (1:4000) was purchased from Thermo Fisher Scientific.

### Cell proliferation and viability assays

Cell proliferation was assessed by using XTT (2, 3–Bis (2–methoxy–4–nitro–5–sulphophenyl)–2H–tetrazolium–5–carboxanilide) (Cat: X12223; Thermo Fisher Scientific Inc.; Waltham, MA, USA), as previously described [16]. Briefly, $Type^{Inv}$ and $Type^{Nec}$ mouse glioblastoma cells were seeded in 96-well plates (4000 cells/ well) in a growth medium with 10% FBS. The cell proliferation was assayed daily through the addition of XTT compound, incubated for 4 h and followed by an absorbance measurement at 450 nM in a BioTek Synergy H1M2 microplate reader (Agilent, Santa Clara, CA, USA).

### Mouse tumor initiation potential

Six-week-old female C57BL/6 mice were injected subcutaneously into left or right flanks with $1 \times 10^6$ mouse brain tumor cells suspended in 100 µL of 25% Matrigel Matrix (BD Biosciences, San Jose, CA, USA) diluted with PBS. Tumor growth was monitored at least twice a week with an electronic digital caliper, and tumor volume was calculated according to the formula: tumor volume (mm3) = (length × breadth × height). Mice were sacrificed at 40 days or when the tumor reached 2000 mm3 in volume.

### Single cell spatial studies and computational typing of conditional mouse glioblastoma model.

CosMx Spatial Molecular Imager (SMI) (Nanostring) was used to detect thousands of RNA transcripts and proteins in formalin-fixed paraffin-embedded (FFPE) spleen tissue sections at subcellular resolution [17]. The CosMx Mouse Universal panels (LBL-11176-03) were utilized. We stained samples with a combination of RNA probes and oligonucleotide conjugated-antibodies that are detected in consecutive rounds of UV-photocleavable reporter hybridization [17]. Tissue brain sections from a mouse tissue microarray were profiled. Each field of view (FOV) captured both transcript coordinates and cell segmentation boundaries at single-cell resolution. CosMx data were collected using the AtoMx software version 3.0.2 (Nanostring),filtered with a custom pipeline, and log library size normalized. Processed transcriptomic and morphology proteomic datasets were analyzed.

**Data Loading and Quality Control**

Raw CosMx flat files (expression matrix, metadata, FOV position, and transcript files) were processed using the Nanostring AtoMx pipeline, and the resulting Seurat object was loaded into R for downstream analysis. Sample-level metadata including organ, histological diagnosis, and GFAP staining status were integrated from the Mouse TMA map. Human xenograft-derived (PDX) samples were excluded for separate analyses, retaining the non-PDX brain tumor cohort. Negative probe controls (negprobes assay) were retained for background assessment. Cells flagged by the AtoMx pipeline were removed. Cells with fewer than 10 detected genes or fewer than 20 total transcripts were excluded.

**Normalization, Batch Correction, and Dimensionality Reduction**

Each sample was normalized independently using SCTransform to account for differences in sequencing depth. To correct for technical variation across samples, Seurat Canonical Correlation Analysis (CCA) integration was performed. Integration anchors were identified using CCA and samples were integrated. Principal component analysis (PCA) was computed on the integrated data. A shared nearest-neighbor graph was constructed on the first 30 principal components, and cells were clustered using the Louvain algorithm. Uniform Manifold Approximation and Projection (UMAP) was computed for visualization.

### Cell Type Annotation

Cluster annotations were established by integrating three independent lines of evidence: (1) marker-based scoring using our curated panel (HES panel) (**Table S8**) of brain cell type markers that we generated derived from the Mouse Whole Brain Atlas publicly available datasets derived from ~4 million cells with a high-resolution transcriptomic and single cell spatial atlas across the mouse brain [18-20]; (2) large language model (LLM) predictions based on cluster-level transcriptomic signatures; and (3) manual curation by domain experts. Final annotations were recorded in a consensus annotation table and applied uniformly across all analyses.

The HES marker panel included canonical markers for major brain cell populations, including dentate gyrus granule cells (*Prox1, Wfs1*), telencephalon inhibitory interneurons (*Sst, Kit*), fibrous astrocytes (*Gfap, Cryab*), radial glia (*Vim, Sox2*), oligodendrocyte precursor cells (OPCs; *Pdgfra, Cspg4, Bmp4*), microglia (*Csf1r, C1qa*), vascular endothelial cells (*Pecam1, Cdh5*), pericytes (*Pdgfrb, Cspg4*), and meningeal/perivascular fibroblasts (*Col1a1, Col3a1, Dcn*). Glioma-associated cell states were identified using markers for OPC-like cells (*Egfr, Pdgfra, Sox9*), proneural glioblastoma (*Erbb3, Pdgfra*), mesenchymal glioblastoma (*Cd44, Fn1, Stat3*), and glioblastoma stem cells (*Sox2, Cd44*).

Our final cell annotation included 22 cell clusters (**Table S7**): Cluster 1 (Cycling OPC-like glioma cells), Cluster 2 (Vascular smooth muscle cells), Cluster 3 (Cycling OPC-like proneuronal glioma cells), Cluster 4 (Activated endothelial cells), Cluster 5 (Glioma associated astrocytes), Cluster 6 (Neuroinflammatory astrocytes), Cluster 7 (Radial glial progenitor cells), Cluster 8 (Glioma associated macrophages/myeloid cells), Cluster 9 (Activated pericytes), Cluster 10 (Vascular leptomeningeal cells (VLMCs) or mural cells), Cluster 11 (Glioma-associated glial Cells), Cluster 12 (Glioma-associated neurons), Cluster 13 (Glioma-associated radial glial cells), Cluster 14 (Glioma-associated microglia), Cluster 15 (Dentate gyrus granule cells), Cluster 16 (Inflammatory neutrophils), Cluster 17 (Inhibitory interneurons), Cluster 18 (Astroependymal cells), Cluster 19 (Choroid plexus epithelial cells), Cluster 20 (Activated neuroinflammatory microglia), Cluster 21 (Meningeal and perivascular fibroblasts), and Cluster 22 (Angiogenic endothelial cells).

### Validation of cell annotation

Annotation validity was independently confirmed against previously reported markers [21-23] and our reference (HES panel) (**Table S8**) based on the Allen Brain Institute (ABI) Mouse Whole Brain Atlas integrating ~4 million cells in a high-resolution transcriptomic and spatial cell type atlas of single cell RNA seq datasets. Key cell annotation were: Oligodendrocyte/Glioma-associated (*Ccnd1, Cd276, Cspg4, Pdgfra*), OPC (*Bmp4, Pdgfra, S100a6*), mature oligodendrocyte/MOL (*Itgb4, Ndrg1, S100b*), astrocyte (*Cd38, Cxcl14, Egfr, Fgfr3, Hgf, Sox9, Vegfa*), microglia (*C1qa, Ccl3, Ccl4, Ccl9, Ccr7, Cd300a, Cd83, Cxcl16, Gdf15, Hck, Il1b, Osm*), endothelial (*Cd34, Efna1, Mecom, Ptgis, Sigirr, Tie1, Vwa1*), pericyte (*Col1a1, Col1a2, Col3a1, Col4a5, Cyp1b1, Dcn, Igf2, S100a10, Twist1, Vtn*), and adult neural stem cell/aNSC (*Birc5, Dll1, Hmgb2, Nusap1, Sox4, Stmn1*).

### $Type^{Inv}$ and $Type^{Nec}$ Glioma Classification

Brain tumors were stratified into Type 1 (classical/NSC lineage) and Type II (proneural/OLC lineage) subtypes based on the expression of previously reported lineage-defining markers [24]. $Type^{Inv}$ markers comprised Egfr and Sox9; the $Type^{Nec}$ marker was Erbb3 (Sox10 was absent from the panel and excluded). A composite $Type^{Inv}$ score was defined as the mean normalized expression of Egfr and Sox9, and a $Type^{Nec}$ score was defined as the normalized expression of Erbb3. Analyses comparing $Type^{Inv}$ and $Type^{Nec}$ expression across histological diagnoses used only pure Type I (Brain tumor (Type I)) and Type II (Brain tumor (Type II)) samples, excluding mixed and GFAP-cre-ERT2 samples where specified.

**Tumor Microenvironment Marker Analysis**

Tumor microenvironment (TME) characterization focused on a curated panel of 11 cytokine, chemokine, and signaling genes associated with macrophage, microglia, and neutrophil activity in the brain tumor context (*Vegfa, Cx3cl1, Vcam1, Igfbp3, Areg, Cxcl2, Cxcl10, Csf1, Ccl5, Hmgb2, Cxcl1/2*). Expression was assessed in core glioma tumor clusters (clusters 1 and 3: Cycling OPC-like glioma cells and Cycling OPC-like proneuronal glioma cells) stratified by histological diagnosis. Immune cell marker expression was further assessed using a panel of macrophage markers (*Itgam, Ptprc, Cd68, Mertk, Cx3cr1, Mrc1, Lyve1, H2-Aa, Tnf, Arg1, Cd163, Ccr2, Cd9, Fcgr4, Cd33, Csf1r, Tlr2, Tlr4, Ccr5, Maf, Pf4, Il1b, S100a9, Cd74, Aif1, Cd86*) and neutrophil markers (*Mpo, Ptprc, Arg1, Cd86, S100a9, Il1b*) across all annotated cell clusters, and in dedicated comparisons between neutrophil, macrophage/myeloid, and microglia populations.

**Differential Gene Expression Analysis**

Differential gene expression (DEG) between cell populations was performed using the Wilcoxon rank-sum test. Genes were considered statistically significant at adjusted $p < 0.05$ (Bonferroni correction) with |average log2 fold change| > 0.25. Two comparisons were performed: (1) Astrocytes versus tumor cells within the GFAP-cre-ERT2 sample; and (2) Glioma-associated glial cells versus Type I/II tumor cells in non-GFAP (ASCL) samples. Results were visualized as volcano plots with −log10(adjusted p-value) on the y-axis and average log2 fold change on the x-axis.

**Spatial Visualization**

For spatial analyses, representative FOVs were selected as the FOV with the greatest number of cells for each histological diagnosis. Cell segmentation polygon boundaries were extracted from the Seurat image slots, which returns polygon vertex coordinates in slide millimeter units. For transcript-level spatial visualization, individual molecule detection coordinates were extracted from the CosMx transcript flat file, which records the FOV-local pixel position and gene identity of each detected transcript. Transcript dot positions were overlaid on cell segmentation polygons, with dot color encoding the per-cell normalized expression level of the corresponding gene. This approach simultaneously displays tissue spatial organization, cell type identity, and within-cell transcript distribution.

**Software and Packages**

All imaging analyses were performed in R (≥ 4.4). Core packages included: Seurat v5 for single-cell data processing, normalization, integration, clustering, and visualization; tidyverse (dplyr, tidyr, ggplot2, purrr) for data manipulation and visualization; pheatmap for heatmap rendering; patchwork for Figurecomposition; viridis for colour scales; ggrepel for non-overlapping gene labels; openxlsx and readxl for Excel import/export; data.table for efficient flat-file parsing; and kableExtra for formatted tables. We utilized the open source Qu-Path for quantitation of necroptosis levels in mouse glioblastoma H&E sections [25].

### Statistical analysis

Data are presented as means ± SEM unless otherwise specified. Seurat FindMarkers with the default Wilcoxon rank-sum test was used for cell marker identification. The Benjamini-Hotchberg procedure was used to make corrections for multiple testing. Survival curves were analyzed using the Kaplan–Meier survival analyses. A P value of <0.05 was used to denote statistical significance unless specifically indicated. Statistical analyses were performed using Prism version 10.0 (GraphPad).

### Data Availability

For MRI image analyses, we retrieved preoperative segmented MRI data from 1,079 glioblastoma patients (585 from the UPENN cohort [8] and 494 from the UCSF cohort [9]). For somatic mutation and expression analyses, we retrieved publicly available mutational data for the AACR Project Genomics Evidence Neoplasia Information Exchange cohort from https://genie.cbioportal.org/. The CNS/Brain datasets at cBioPortal included 3,639 glioblastoma patients from the Diffuse Glioma (GLASS Consortium) (n=329 patients) [26], Diffuse Glioma (GLASS Consortium, 2019) (n=222 patients) [27], Diffuse Glioma (TCGA, GDC, 2025) (n=516 patients) [28], Glioblastoma (TCGA, 2013) (n=577 patients) [29], Glioblastoma (TCGA, 2008) (n=206 patients) [30], Glioblastoma multiforme (TCGA, 2025) (n=598 patients) [28], Glioblastoma multiforme (TCGA, Legacy) (n=606 patients) [31], and Glioblastoma multiforme (Pan-Cancer Atlas) (n=585 patients) [32]. For glioblastoma mouse brain single cell spatial cell typing, we retrieved high-resolution transcriptomic and spatial cell type atlas of single cell RNA seq datasets from the Allen Brain Institute (ABI) Mouse Whole Brain Atlas integrating ~4 million cells to generate references for clustering from previously reported normal brain cell type markers [21-23].

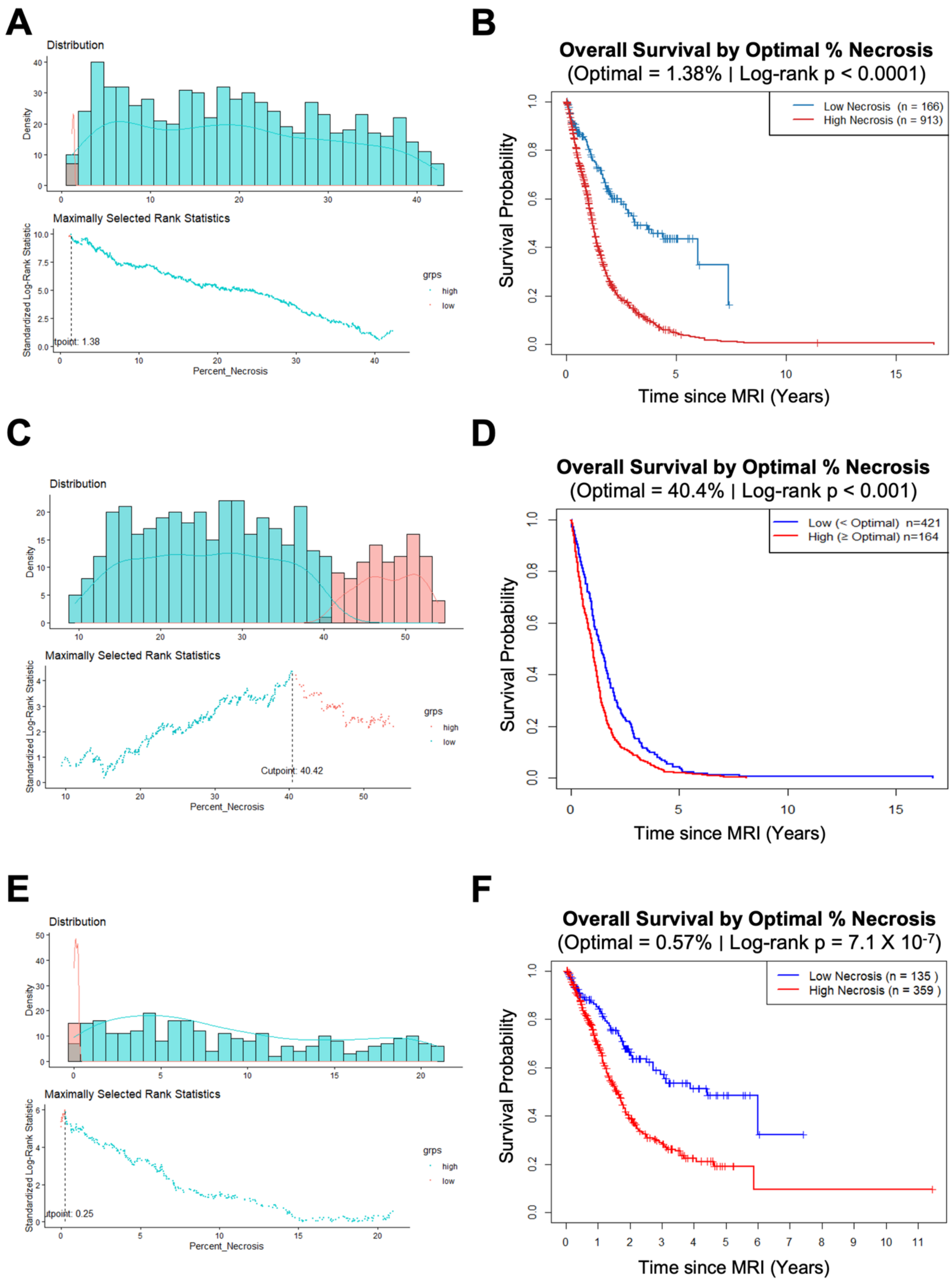


**Figure S1**. **High tumor necrosis among heterogeneous glioblastoma patients and associated with worse survival.** (**A**) Necrosis density distribution (Top) and maximally selected ranked statistics (Bottom) to identify the optimal cut off (tpoint, 1.38) of the percent necrosis between the two groups of high and low necrosis within both cohorts combined. (**B**) Kaplan–Meier survival curves stratified by statistically determined optimal necrotic core percentage (1.38%) in patients with diffuse glioblastoma from the UPENN and UCSF cohort (n = 1,079). Patients with high necrosis (≥1.38%, red, n=913) demonstrated significantly shorter overall survival compared with those with low necrosis (<1.38%, blue, n=539). Log-rank test, P < 0.0001. (**C**) Necrosis density distribution (Top) and maximally selected ranked statistics (Bottom) to identify the optimal cut off (tpoint, 40.4%) within the UPENN cohort. (**D**) Kaplan–Meier survival curves stratified by statistically determined optimal necrotic core percentage (40.4%) in the UPENN cohort (n = 585). Patients with high necrosis (≥40.4%, red, n=164) demonstrated significantly shorter overall survival compared with those with relatively low necrosis (<40.4%, blue, n=421). Log-rank test, P < 0.001. (**E**) Necrosis density distribution (Top) and maximally selected ranked statistics (Bottom) to identify the optimal cut off (tpoint, 0.57%) within the UCSF cohort. (**F**) Kaplan–Meier survival curves stratified by statistically determined optimal necrotic core percentage (0.57%) in the UCSF cohort (n = 494). Patients with high necrosis (≥0.57%, red, n= 359) demonstrated significantly shorter overall survival *vs.* those with relatively low necrosis (<0.57%, blue, n=135). Logrank test, $P = 7.1 \times 10^{-7}$.

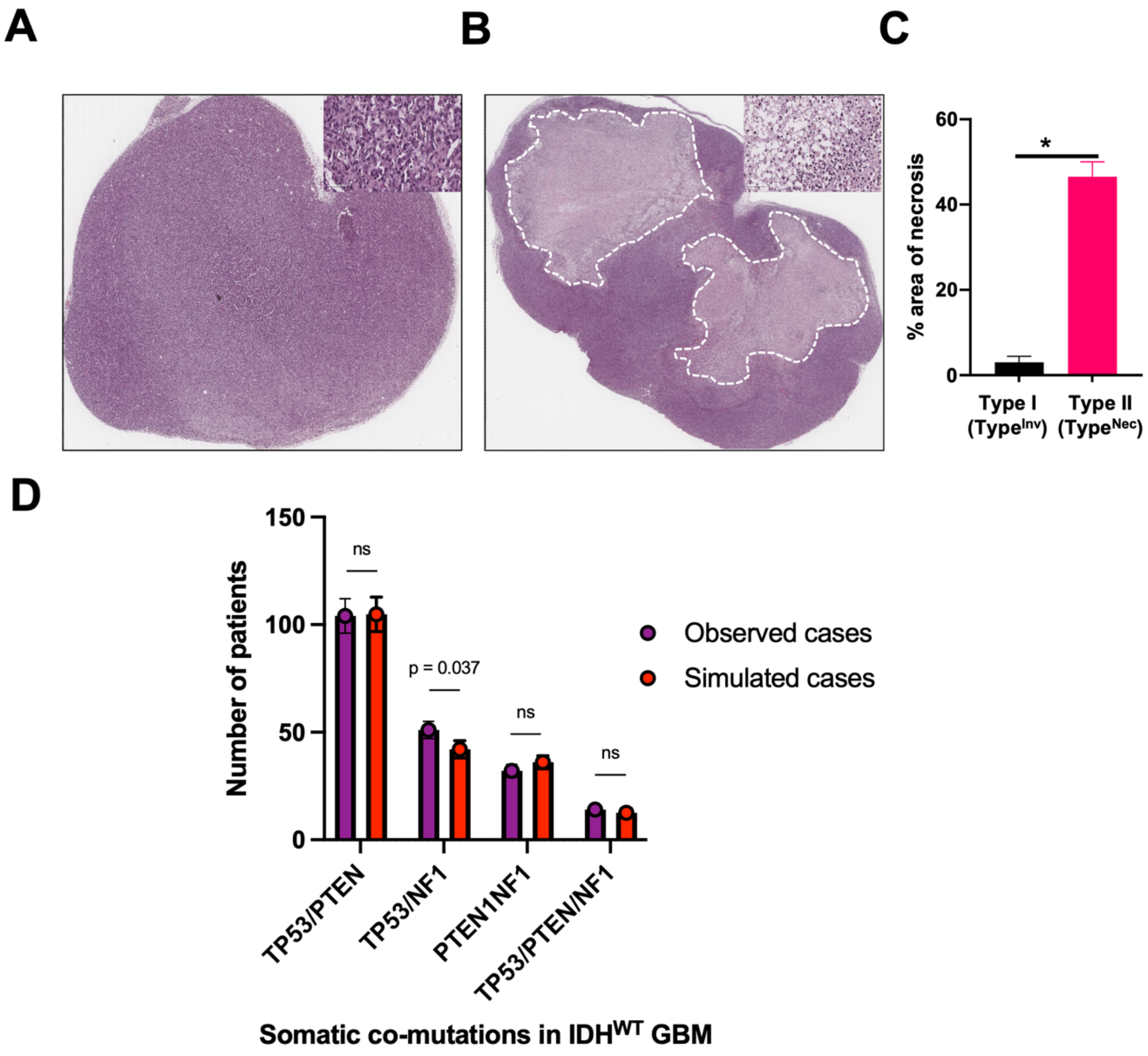


**Figure S2. Human glioblastoma with PPN somatic mutations propagate the necroptotic phenotype in orthotopic mouse xenografts.** (**A-B**) H&E sections of representative orthotopic mouse xenografts (n=10) from glioblastoma patient derived organoids (PDOs) [14], from an original patient glioblastoma tissues H&E sections either not demonstrating necrosis (**A**) or demonstrating pseudopalisading necrosis (**B**). Insets are higher magnifications from the outlined areas. (**C**) Quantitation of necroptosis levels in mouse glioblastoma sections using Qu-Path software. (**D**) *TP53, PTEN, NF1* (PPN) somatic mutation multi-gene permutation model. Monte Carlo simulations with comparison of the observed clinical cohort count against the simulated null distribution (p-value = 0.03785).

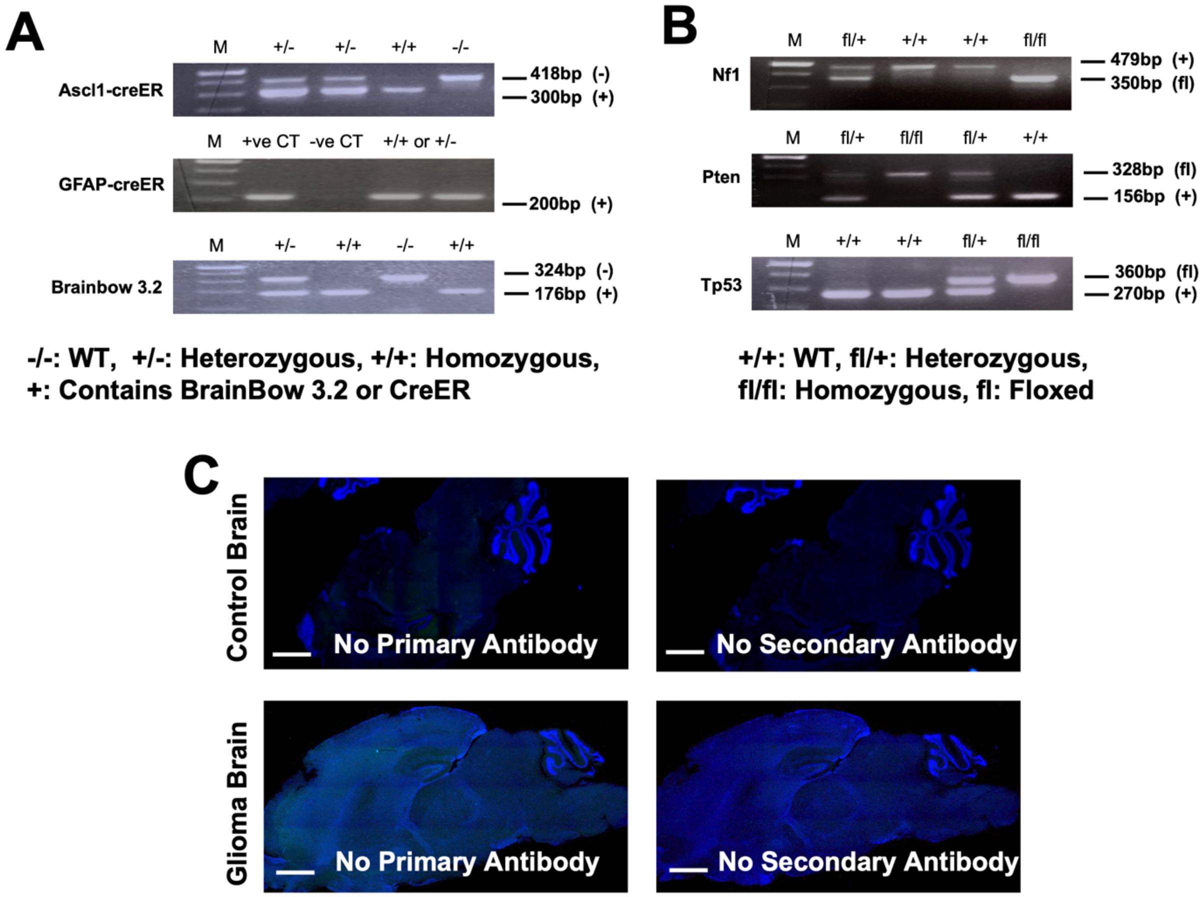


**Figure S3. Genotyping of the ANPPB compound GEMMM of glioblastomas.** (**A**) Representative genotyping of cre-ER and Brainbow fluorescent reporters. (**B**) Genotyping of *NF1, PTEN*, and *TP53* floxed tumor suppressors to generate conditional compound loss of *NF1, PTEN,* and *TP53* in Ascl1 and GFAP-expressing brain cells. (**C**) Control IF staining of the Brainbow fluorescent variant with no signal detected when primary or secondary antibodies are omitted.

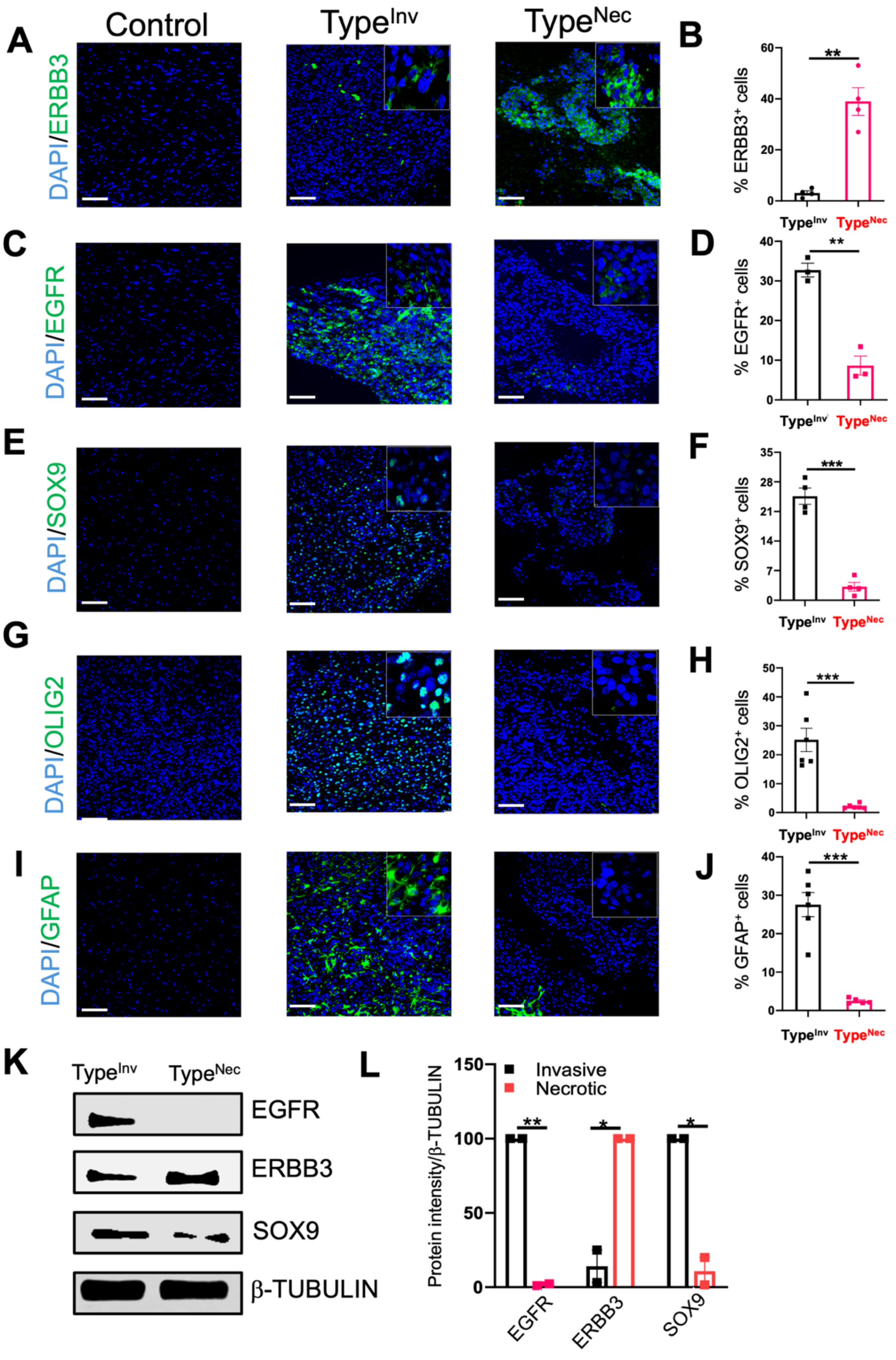


**Figure S4**. **Distinguishable molecular signatures of invasive (Type$^{Inv}$) and necrotic (Type$^{Nec}$) glioblastomas.** (**A-J**) Immuno-fluorescence (IF) analysis of ERBB3 (**A**), EGFR (**C**),

SOX9 (**E**), OLIG2 (**G**), and GFAP (**I**) expression (green) in control brain (left), invasive (center), and necrotic tumor section (right), and representative histogram of intensity of ERBB3 (**B**), EGFR (**D**), SOX9 (**F**), OLIG2 (**H**), and GFAP (**J**) in glioblastomas. (**K**) WB of ERBB3, SOX9, and EGFR expression in invasive and necrotic glioblastoma cell lines, normalized based on equal loading from similar cell numbers by β-TUBULIN expression. (**L**) Representative histogram of band intensity of EGFR, ERBB3, and SOX9 in respective groups in the study. Representative data are shown and expressed as the mean ± SEM, *$p < 0.05$, ** $p < 0.01$, and *** $p < 0.001$. Scale bars are 100 μm.

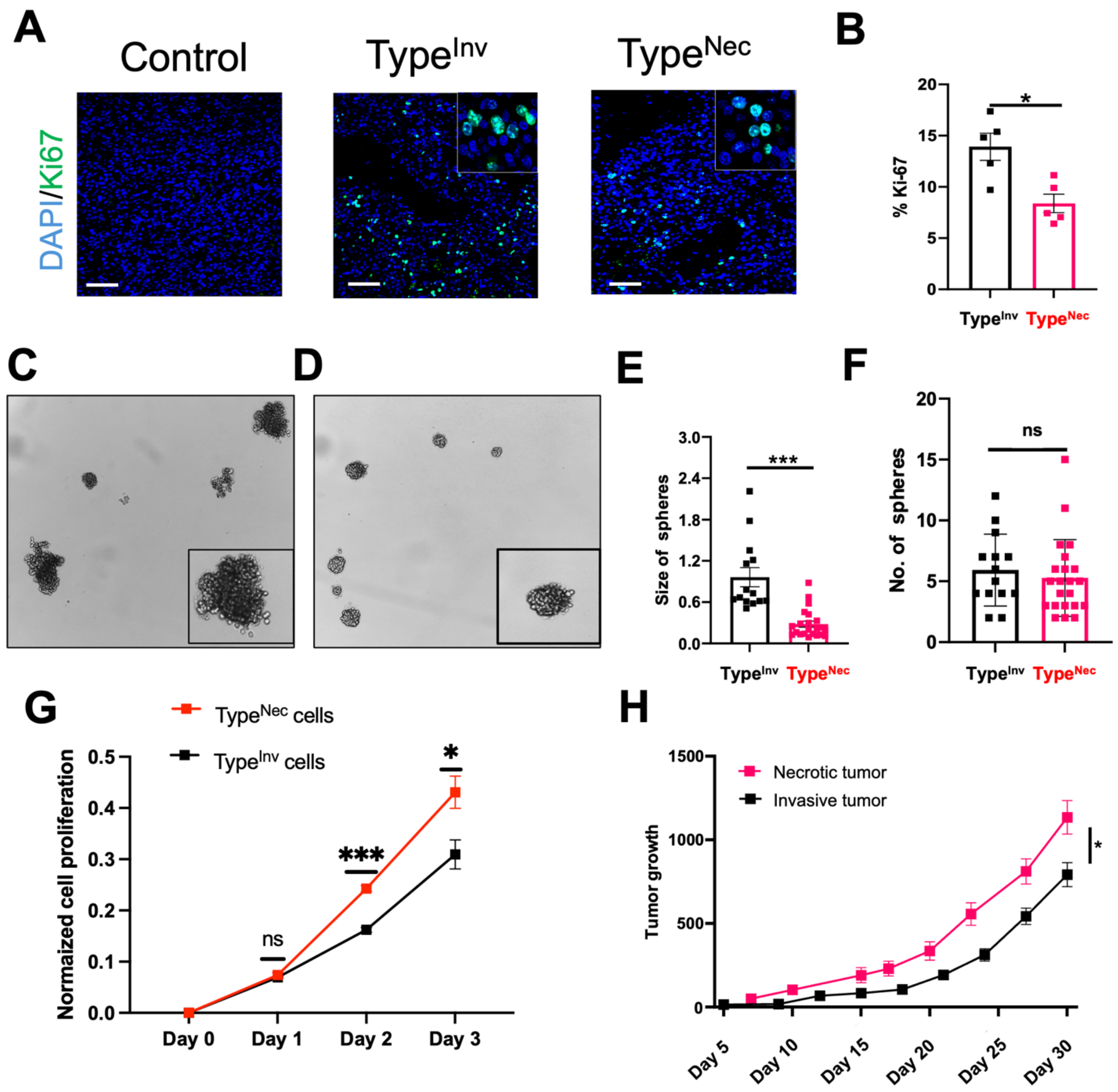


**Figure S5**. **Distinguishable phenotypes of invasive tumor and necroptotic glioblastomas.** (**A-D**) Characterization of molecular signatures in the tumor tissue. (**A**) Immuno- fluorescence analysis of Ki-67 in invasive tumor section (left), necrotic tumor section (center) and control brain tissue (right). (**B**) Representative histogram of intensity of Ki-67 in tumors of respective tumor groups in the study. (**C-F**) Analysis of functional studies between invasive and necrotic glioblastoma cell lines. Tumor-sphere-forming assay was conducted in 6-well plates to assess the self-renewal ability of invasive (**C**) and necroptotic (**D**) glioblastoma cell lines. (**E**) Representative histogram of size of tumor-spheres formed in the invasive (n=10) and necrotic (n=10) glioblastoma cells. (**F**) Representative histogram of number of tumor-spheres formed in the invasive (n=10) and necrotic (n=10) glioblastoma cells. (**G**) Cell proliferation assay of invasive and necrotic glioblastoma cell lines was assessed with XTT assay. (**H**) Isolated mouse glioblastoma cells ($1x10^6$ of necroptotic tumor cells (E2115)) or ($1x10^6$ of invasive tumor cells (E2203)) were injected subcutaneously into 6 weeks old C57BL6 mice. Tumor volume was measured using a caliper twice a week. Representative data are shown and expressed as the mean ± SEM (n=3), *$p < 0.05$, ** $p < 0.01$, and *** $p < 0.001$. Scale bars are 100 μm.

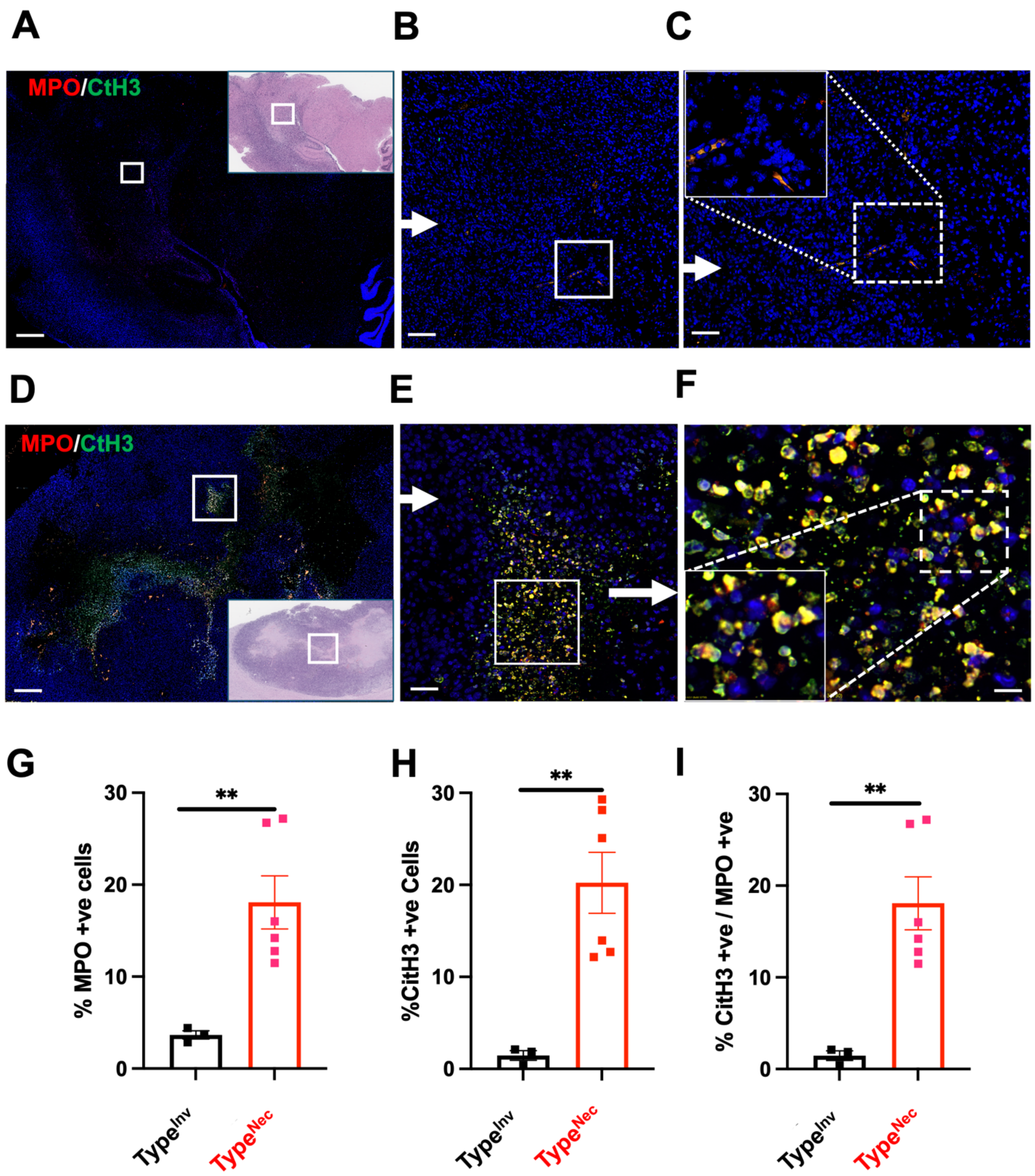


**Figure S6**. **Necrotic tumor regions are highly enriched with neutrophil extracellular traps (NETs).** (**A-C**) Immune fluorescence staining of MPO-(red), Citrullinated histone H3 (CitH3) -(green) and DAPI (blue) in the whole invasive tumor tissue with 10x magnification (**A**), with the H&E inset of the corresponding tumor region. (**B-C**) The same region stained in invasive tumor tissue with MPO (red), CitH3 (green) and DAPI in higher magnification, and cellular localization of staining shown in the insert (**C- top left corner**).

(**D-F**) Immune fluorescence staining of MPO-(red), Citrullinated histone H3 (CitH3) -(green) and DAPI (blue) in necrotic glioblastoma (**D**), with the H&E inset of the corresponding tumor region. (**E-F**) The same stained region of necrotic tumor tissue with MPO (red), CitH3 (green) and DAPI with higher magnification, and cellular localization of the staining shown in the insert (**C**- top left corner). (**G**) Representative histogram of percentage of double positive (MPO-red and CitH3-green) in the invasive (n=3) and necrotic (n=3) glioblastoma tumor tissue. Representative data expressed as the mean ± SEM (n =3), ** $p < 0.01$. Scale bars are 1 mm in **A** and **D**, and 100 μm in **B**, **C**, **E**, and **F**.

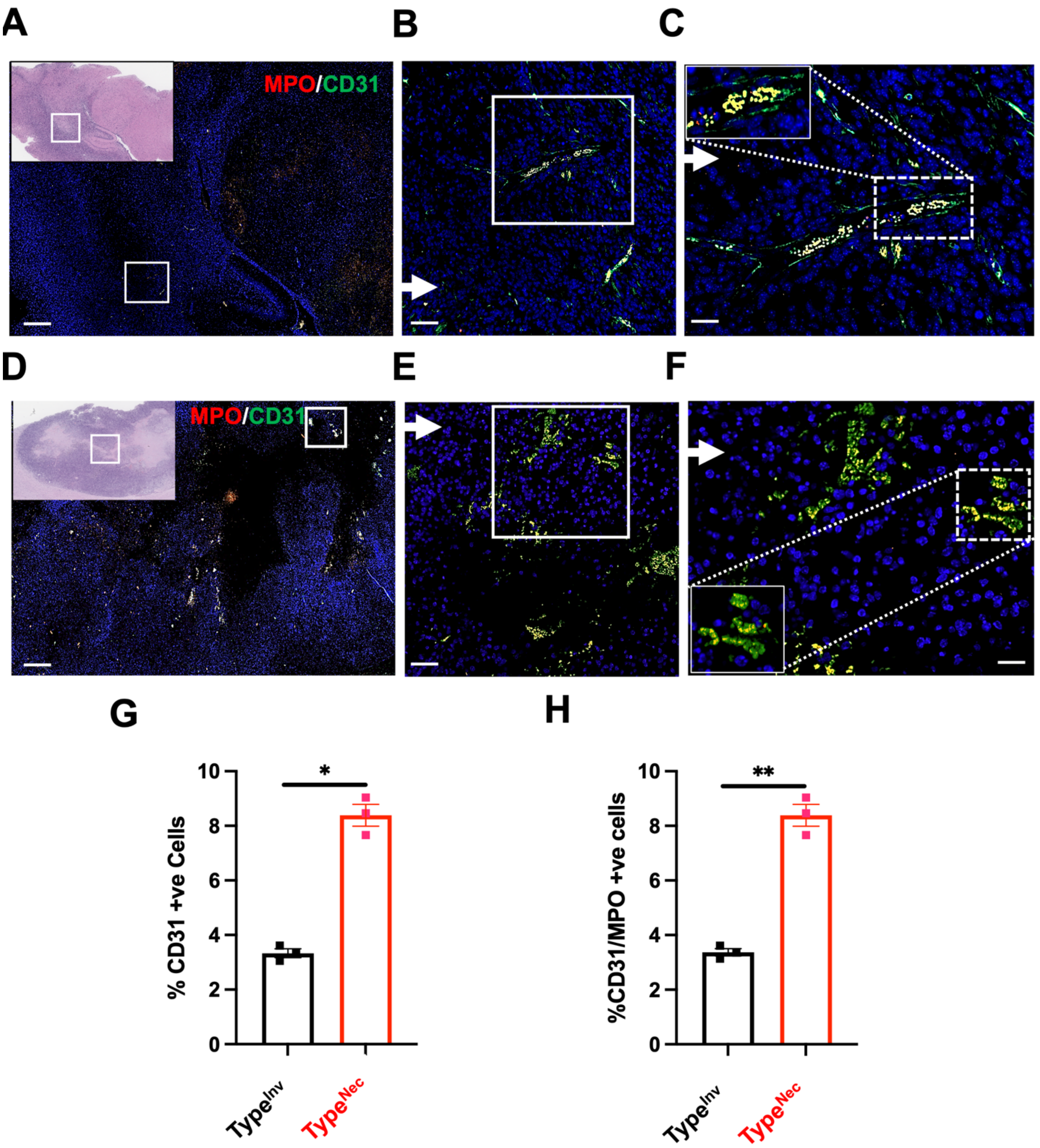


**Figure S7**. **Necroptotic tumor tissue is rich in neutrophils present in blood vessels.** (**A-C**) Immune fluorescence staining of MPO-(red), CD31-(green) and DAPI (blue) in the whole invasive tumor tissue with 10x magnification (**A**), the H&E staining of the corresponding tumor tissue (in insert, top-left). (**B-C**) The stained invasive tumor tissue with MPO (red), CD31 (green) and DAPI with higher magnification, and cellular localization of staining shown in the insert (**C**- top left corner). (**D-F**) Immune fluorescence staining of MPO-(red),

CD31-(green) and DAPI (blue) in the whole necrotic tumor tissue with 10x magnification (**D**), the H&E staining of the corresponding tumor tissue (in insert, bottom-left). Note the $CD31^{+}$ highly abnormal, disorganized glomeruloid vascular structures, a definitive histopathological hallmark of glioblastoma. (**E-F**) The stained necrotic tumor tissue with MPO (red), CitH3 (green) and DAPI with higher magnification, and cellular localization of staining shown in the insert (**C**- top left corner). (**G-H**) Representative histogram of percentage of single (**G**) and double positive (MPO-red and CD31-green) (**H**) in the invasive (n=3) and necrotic (n=3) glioblastoma tumor tissue. Representative data are shown and expressed as the mean ± SEM (n =3), *$p < 0.05$, ** $p < 0.01$, and *** $p < 0.001$. Scale bars are 1 mm in **A** and **D**, and 100 μm in **B**, **C**, **E**, and **F**.

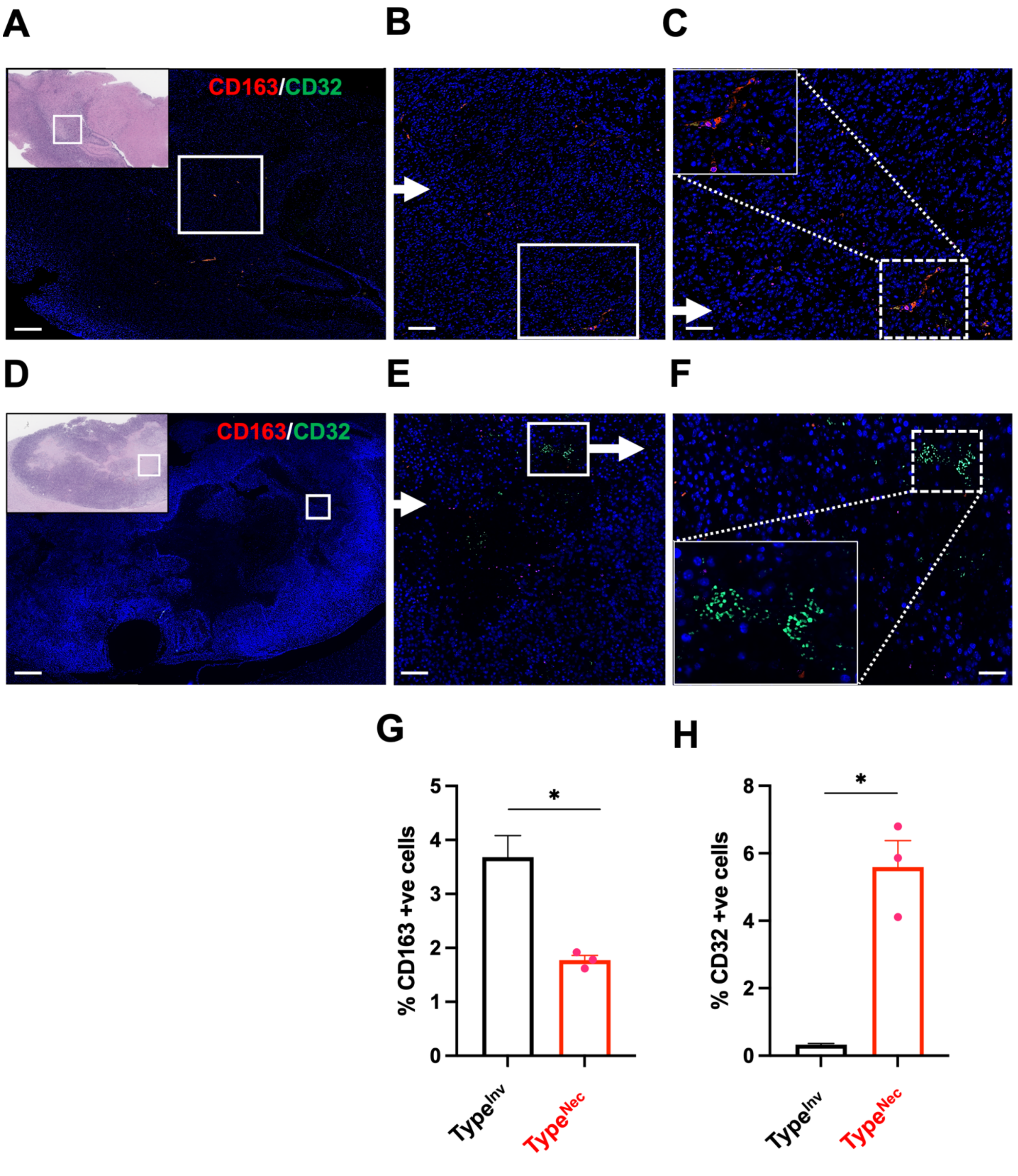


**Figure S8**. **Distribution of pro-inflammatory and anti-inflammatory macrophages in invasive *vs.* necrotic glioblastomas.** (**A-C**) Immune fluorescence staining of CD163-(red), CD32 -(green) and DAPI (blue) in the whole invasive tumor tissue with 10x magnification (**A**), H&E staining of the corresponding tumor tissue (in insert, top-right). (**B-C**) The stained invasive tumor tissue with CD163 (red), CD32 (green) and DAPI with higher magnification, and cellular localization of staining shown in the insert (**C**- bottom right corner). (**D-F**) Immune fluorescence staining of Cd163-(red), CD32-(green) and DAPI (blue) in the whole necrotic tumor tissue with 10x magnification (**D**), the H&E staining of the corresponding tumor tissue (in insert, top-right). (**E-F**) The stained necrotic tumor tissue with CD163 (red), CD32 (green) and DAPI with higher magnification, and cellular localization of staining shown in the insert (**C**- top right corner). (**G-H**) Representative histogram of percentage of positive of CD163 (red) (**G**) and CD32 (green) (**H**) in the invasive (n=3) and necrotic (n=3) glioblastoma tumor tissue. Representative data are shown and expressed as the mean ± SEM (n =3), $*p < 0.05$, $** p < 0.01$, and $*** p < 0.001$. Scale bars are 500 μm in **A** and **D**, and 100 μm in **B**, **C**, **E**, and **F**.

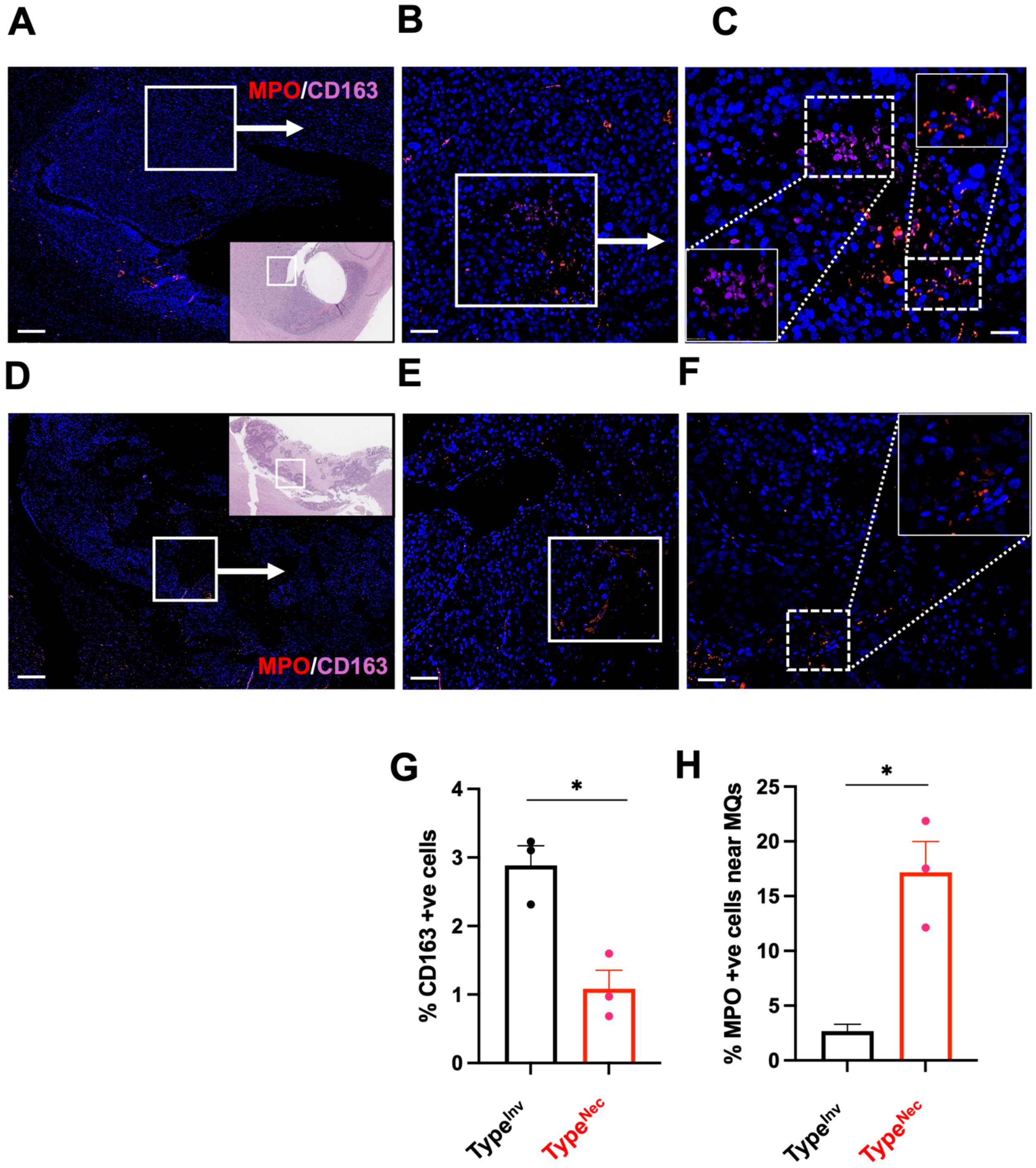


**Figure S9**. **Presence of anti-inflammatory macrophages promotes the necroptosis in the glioblastoma brain tumor tissue.** (**A-C**) Immune fluorescence staining of MPO-(red), CD163 -(purple) and DAPI (blue) in the whole invasive tumor tissue with 10x magnification (**A**), the H&E staining of the corresponding tumor tissue (in insert, top-right). (**B-C**) The stained invasive tumor tissue with MPO (red), CD163 (purple) and DAPI with higher magnification, and cellular localization of staining shown in the insert (**C**-bottom right corner). (**D-F**) Immune fluorescence staining of MPO-(red), CD163-(purple) and DAPI (blue) in the whole necrotic tumor tissue with 10x magnification (**D**), the H&E staining of the corresponding tumor tissue (in insert, top-right). (**E-F**) The stained necrotic tumor tissue with MPO (red), CD163 (green) and DAPI with higher magnification, and cellular localization of staining shown in the insert (**C**- top right corner). (**G-H**) Representative histogram of percentage of cells positive for CD163 (purple) (**G**) and $MPO^+$ cells within 50 μm from $CD163^+$ cells (red) (**H**) in the invasive (n=3) and necrotic (n=3) glioblastoma tumor tissue. Representative data are shown and expressed as the mean ± SEM (n=3), *$p < 0.05$, ** $p < 0.01$, and *** $p < 0.001$. Scale bars are 500 μm in **A** and **D**, and 100 μm in **B**, **C**, **E**, and **F**.

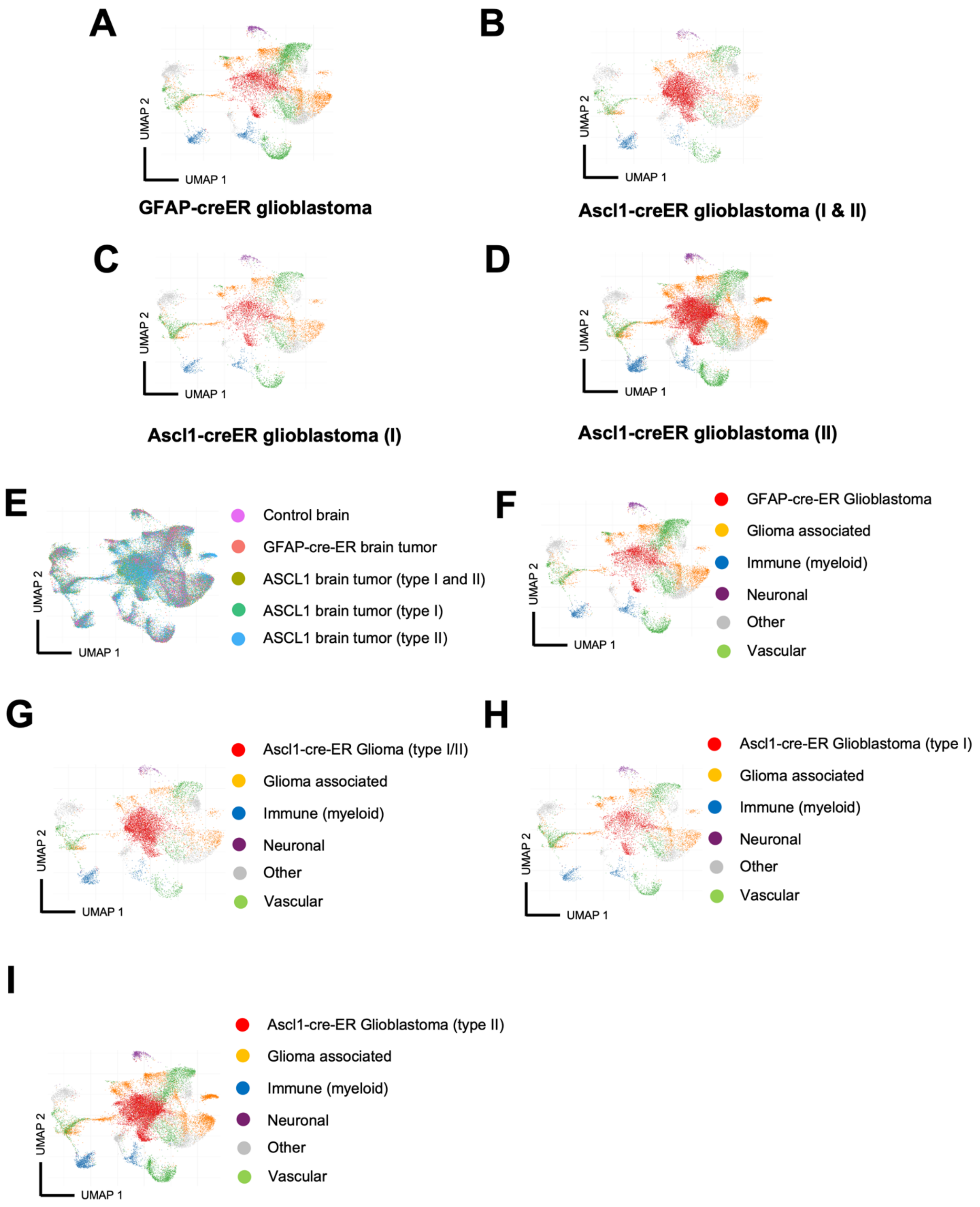


**Figure S10. Single cell spatial visualization of glioblastoma tumor cells and associated TME.** (**A-D**) UMAP visualization of mouse glioblastoma tumor cells and TME associated annotated clusters and proportions of the 22 mouse brain cell clusters (listed in **Table S7**)

across samples, colored by cell type-annotation across mouse brain sections from GFAP-cre-ER (**A**), Ascl1-cre-ER type I (Type$^{Inv}$) and II (Type$^{Nec}$) (**B**), type I only (**C**), and type II (Type$^{Nec}$) only (**D**) mouse glioblastomas. (**E-I**) UMAP visualization of detailed cell types of mouse glioblastoma tumor cells and TME associated, immune, myeloid, neuronal, vascular, and other cells colored by cell type-annotation across mouse brain sections from various tissue (**E**), GFAP-cre-ER (**F**), type I and II (**G**), type I only (**H**), and type II only (**I**) mouse glioblastomas.

**A**

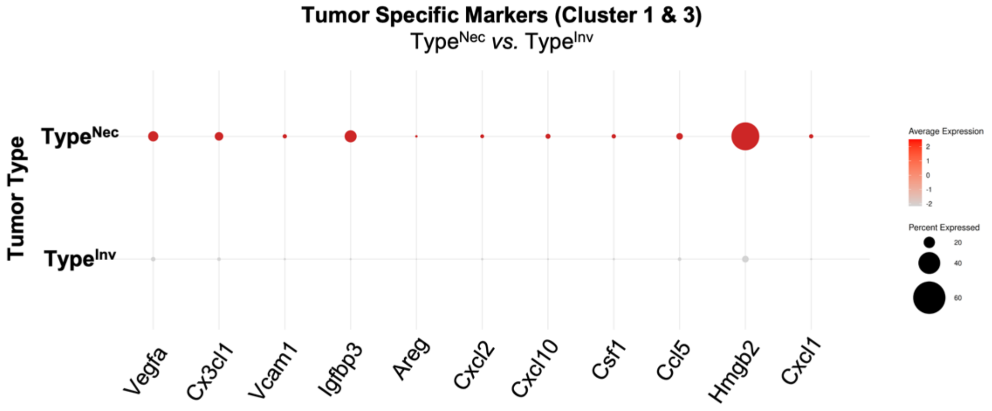


**B**

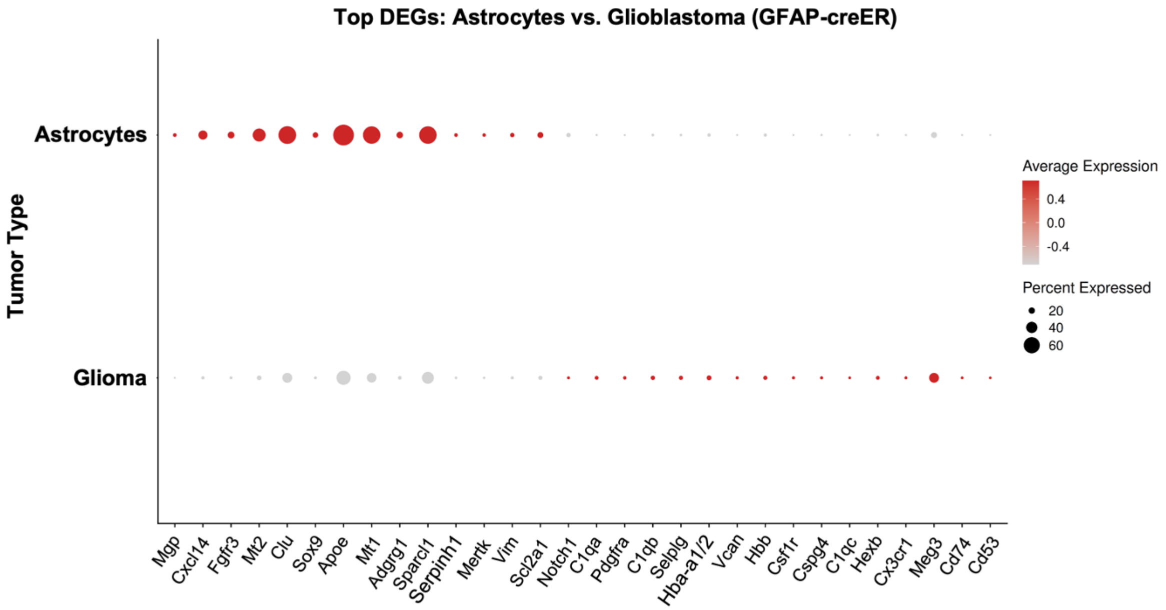


**Figure S11. Comparison of glioblastoma tumor cell specific markers between subtypes.** (**A**) Bubble heatmap comparing the expression levels of select marker genes in tumor specific clusters 1 and 3 comparing type I ($Type^{Inv}$) and II ($Type^{Nec}$) glioblastomas. (**B**) Bubble heatmap showing the expression levels of select marker genes comparing the expression levels of select marker genes in tumor specific GFAR-creER *vs.* astrocytes as potential tumor initiating cells. Color indicates normalized average expression, and dot size indicates the percentage of expressing cells.

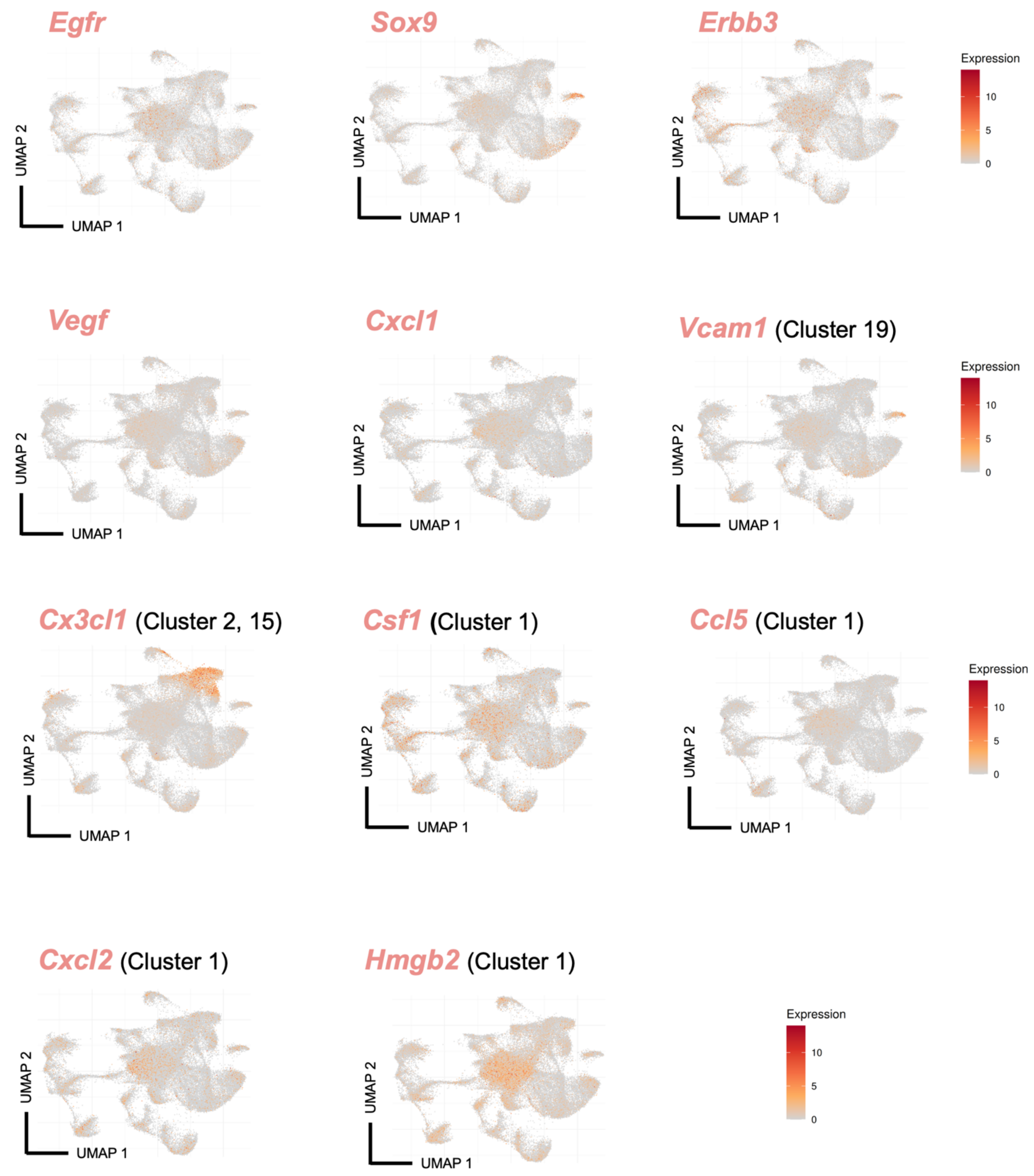


**Figure S12. Cell type specific transcripts in single cell spatial mapping of the glioblastoma TME.** Transcript UMAP visualization of and expression levels of TME associated 22 mouse brain cell clusters. Expression levels and cluster specific expression of *EGFR, SOX9, ERBB3, VEGF, CXCL1, CSF1, CCL5, CXCL2*, and *Hmgb2* predominantly in cluster 1 and 3 (tumorigenic) cells. *VCAM1* is predominantly expressed in choroid plexus in epithelial cells, while *CX3CL1* enriched in VSMCs and Dentate gurus granule cells among other cell clusters.

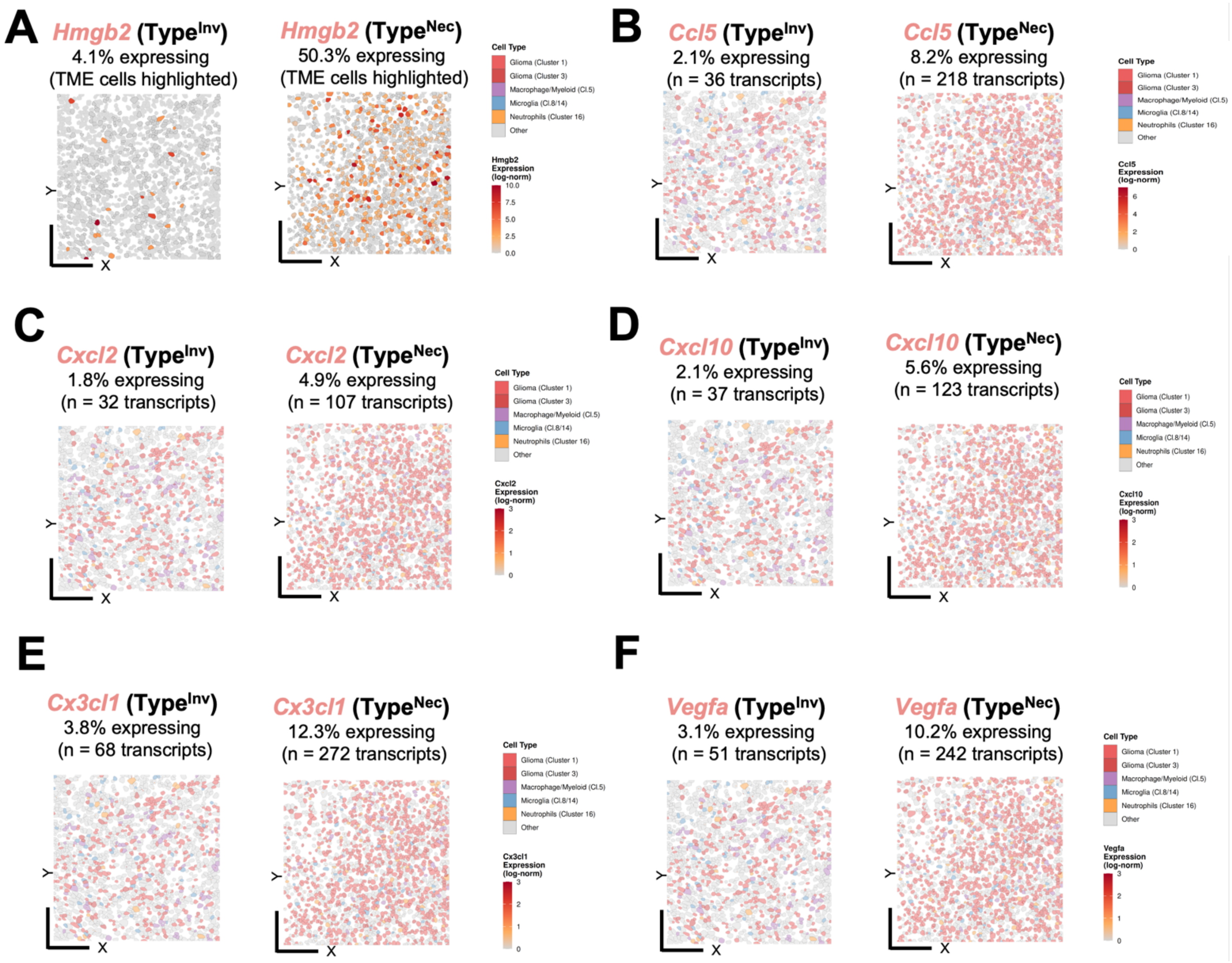


**Figure S13**. **Glioma subtype specific markers in single cell spatial mapping TME.** (**A-F**) Single cell spatial cell type map from type I (Type$^{Inv}$) *vs.* type II (Type$^{Nec}$) glioblastoma colored by cell clusters of glioblastoma cells and localized RNA transcripts. Percentage of cells expressing, transcript levels in single cells and cell type specific expression of *Hmgb2* (**A**), *Ccl5* (**B**), *Cxcl2* (**C**), *Cxcl10* (**D**), *Cx3cl1* (**E**), and *Vegfa* (**F**). Data represent 59,099 single cell spatial data from 16 mouse brain sections.

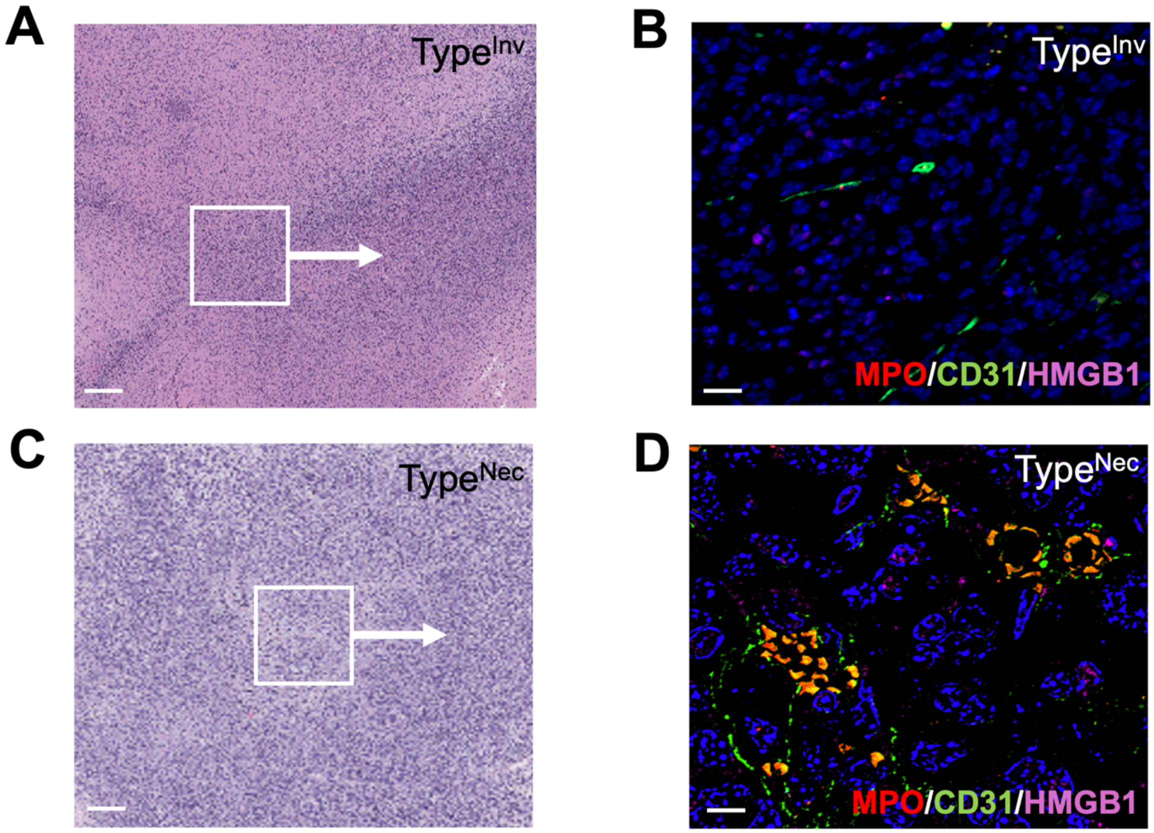


**Figure S14**. **Identification of DAMPs in necroptotic and perivascular regions of Type$^{Nec}$ glioblastomas enriched with NETs.** (**A-D**) H&E and immune fluorescence staining of MPO-(red), CD31(green) and HMGB1 (purple) in type I (Type$^{Inv}$) (**A-B**), and type II (Type$^{Nec}$) (**C-D**). Scale bars are 1 mm in **A** and **C**, and 100 μm in **B** and **D**.

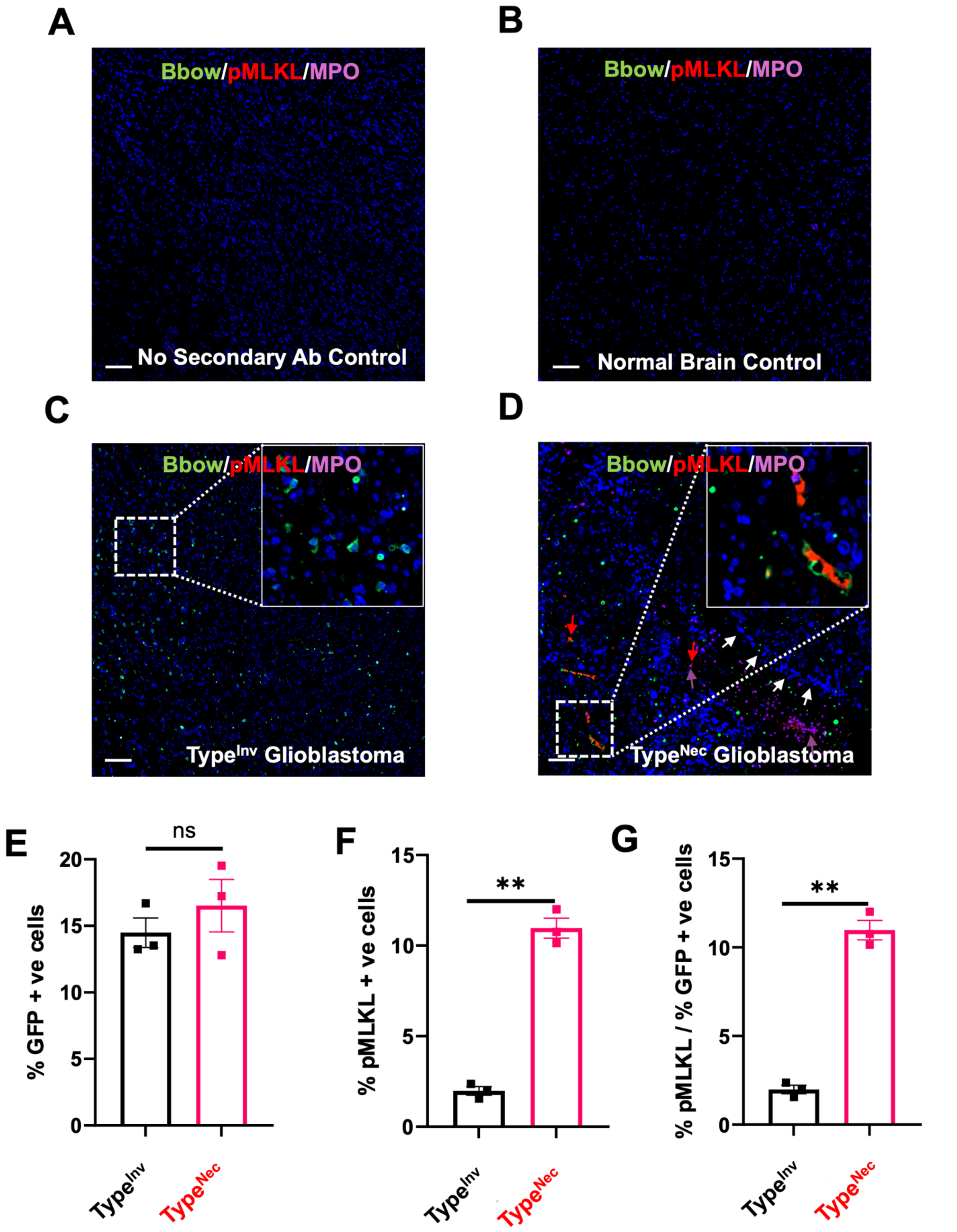


**Figure S15**. **DAMP signaling and activation of necroptosis in the necroptotic glioblastoma.** (**A-D**) Control immune fluorescence staining of phosphorylated MLKL (pMLKL) as the necroptosis executer, EGFP variant for the glioblastoma tumor cell derived Brainbow (Bbow)-green, MPO-(purple) for NETs, and DAPI (blue) in antibody control (omitting secondary antibody (**A**), normal mouse brain (**B**), invasive tumor tissue (**C**), and necroptotic glioblastoma (**D**), with higher magnification and cellular localization of staining shown in the insert (**C-D** upper right corner). White arrows indicate pseudopalisading necrosis, purple arrows are for NETs, and the red arrow points to cell membrane pMLKL. (**E-F**) Representative histogram of the percentage of cells positive for variant GFP for Brainbow (green) (Glioblastoma tumor cells) (**E**) and pMLKL$^+$ cells within 50 μm from MPO$^+$ cells (purple) (**F**) in the invasive (n=3) and necrotic (n=3) glioblastoma tumor tissue. (**G**) The ratio of pMLKL necroptosis executer over GFP-positive glioblastoma tumor cells in invasive *vs.* necroptotic glioblastomas. Representative data are shown and expressed as the mean ± SEM (n=3), $*p < 0.05$, $** p < 0.01$, and $*** p < 0.001$. Scale bars are 100 μm in **A-D**.

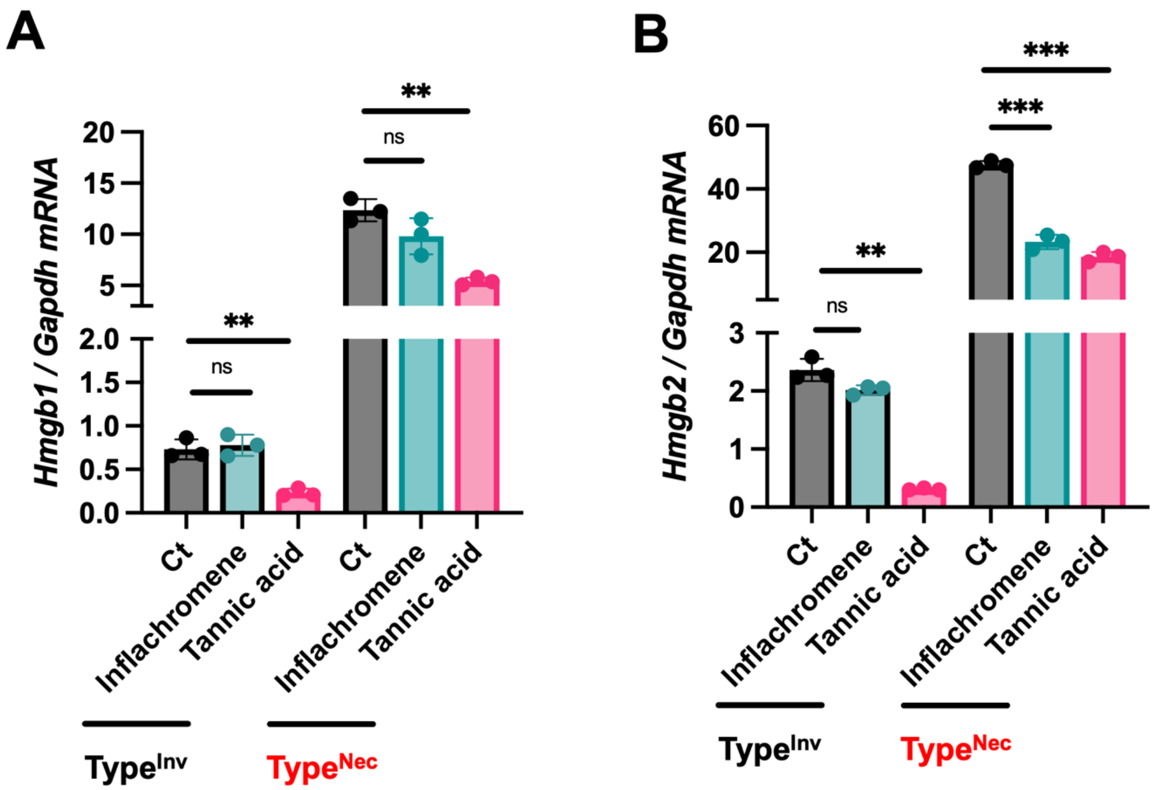


**Figure S16**. **Pharmacological suppression of DAMP transcriptome using anti-inflammatory compounds.** (**A-B**) Anti-inflammatory drugs, Inflachromene and Tannic acid target and significantly reduce *Hmgb1* and *Hmgb2* DAMP transcripts in glioblastoma cells, relative to vehicle control (Ct), supporting a potential combination therapeutic strategy with standard of care antiproliferative agents. These anti-inflammatory agents (Inflachromene is an experimental neuroinflammatory and microglia inhibitor, while Tannic acid is an FDA approved safe food additive) have no direct antiproliferative activity. Representative data are shown and expressed as the mean ± SEM (n = 3 experiments), ** $p < 0.01$.

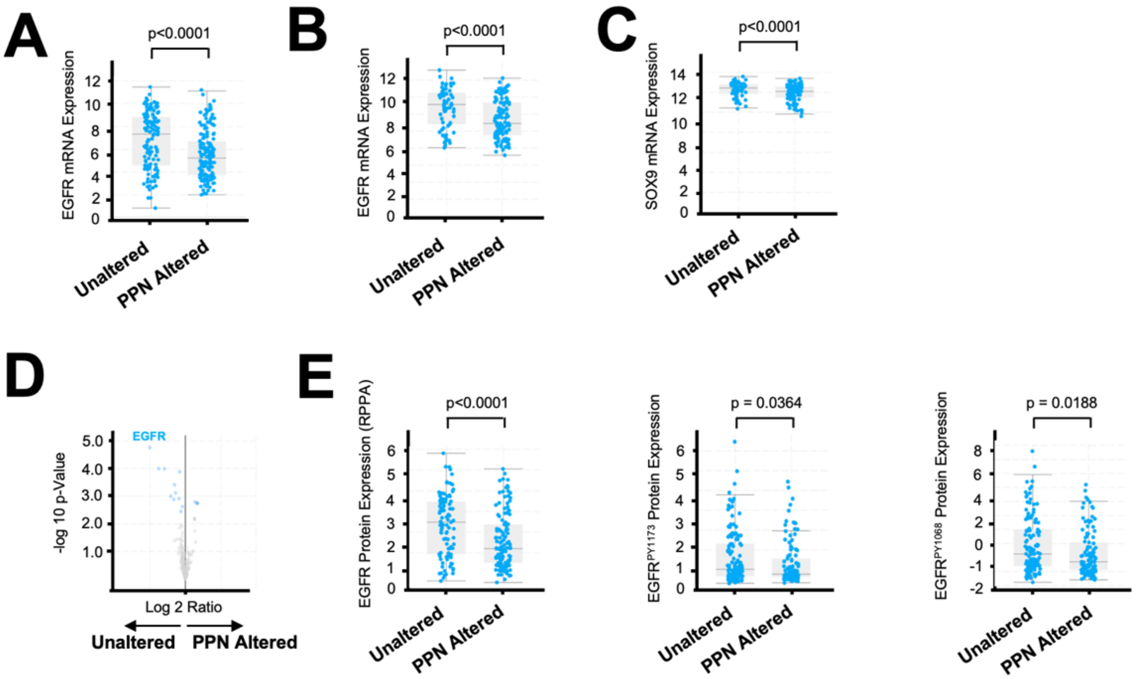


**Figure S17**. **Higher EGFR expression associated with PPN unaltered type I (Type$^{Inv}$) human glioblastoma**. (**A-C**) Box correlation plot showing *EGFR* mRNA expression among the TCGA GDC (A) and legacy (**B**) datasets, and significantly higher *EGFR* and *SOX9* (**C**) expression in PPN unaltered (n=286 patients) *vs.* PPN altered (n=237 patients). **(D)** Protein expression was only available among the glioblastoma patients within the TCGA Legacy dataset. Volcano plot showing differentially expressed proteins (DEPs) between PPN unaltered and altered samples, with EGFR protein highest DEP associated with PPN unaltered *vs.* PPN altered. (**E**) Additional plots showing significantly higher EGFR$^{PY1173}$ and EGFR$^{PY1068}$ proteins with PPN unaltered *vs.* PPN altered Two-tailed unpaired t-test were used for statistical comparisons of correlates.

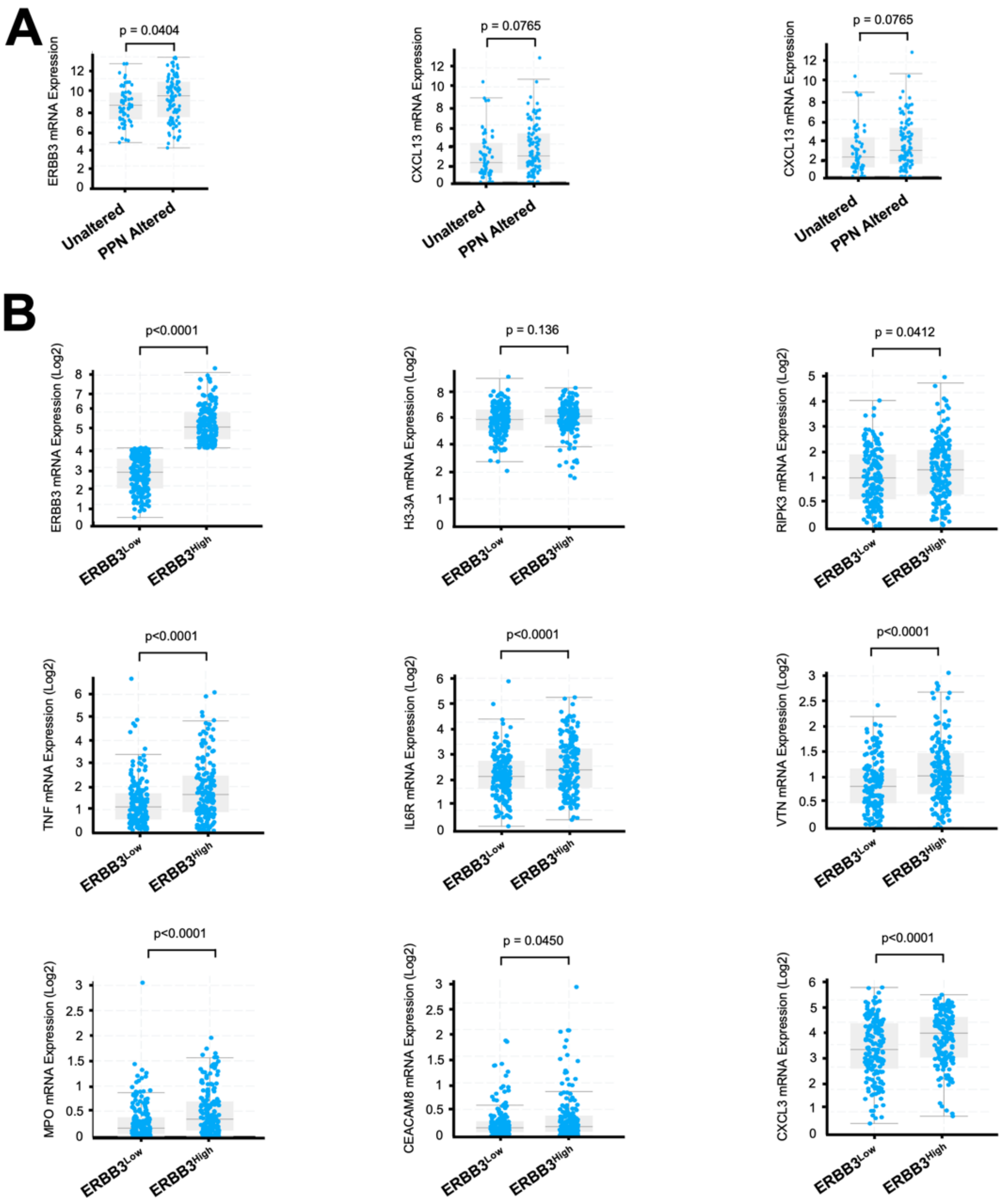


**Figure S18**. **ERBB3 expression associated with PPN myeloid immunoinflammatory niche in human glioblastoma patients**. (**A**) Box correlation plot showing *ERBB3* mRNA expression among the TCGA legacy dataset (n=591 glioblastoma patients), and significantly higher *ERBB3* expression in PPN altered (n=213 patients) *vs.* PPN unaltered (n=378 patients). **(B) C**orrelation plot of mRNA expression (Log2 score) among glioblastoma patients within the GLASS consortium [26] comparing the transcriptomic signature of samples with *ERBB3*$^{High}$ (n=121 patients) *vs. ERBB3*$^{Low}$ (n=121 patients) for the histone *H3-3A*, DAMP activation (*RIPK3*), inflammatory recruitment (*TNF, IL6R*, and *VTN*), and myeloid-derived activation (*MPO, CEACAM8*) with *CXCL3*. Two-tailed unpaired t-test were used for statistical comparisons of mRNA correlates.

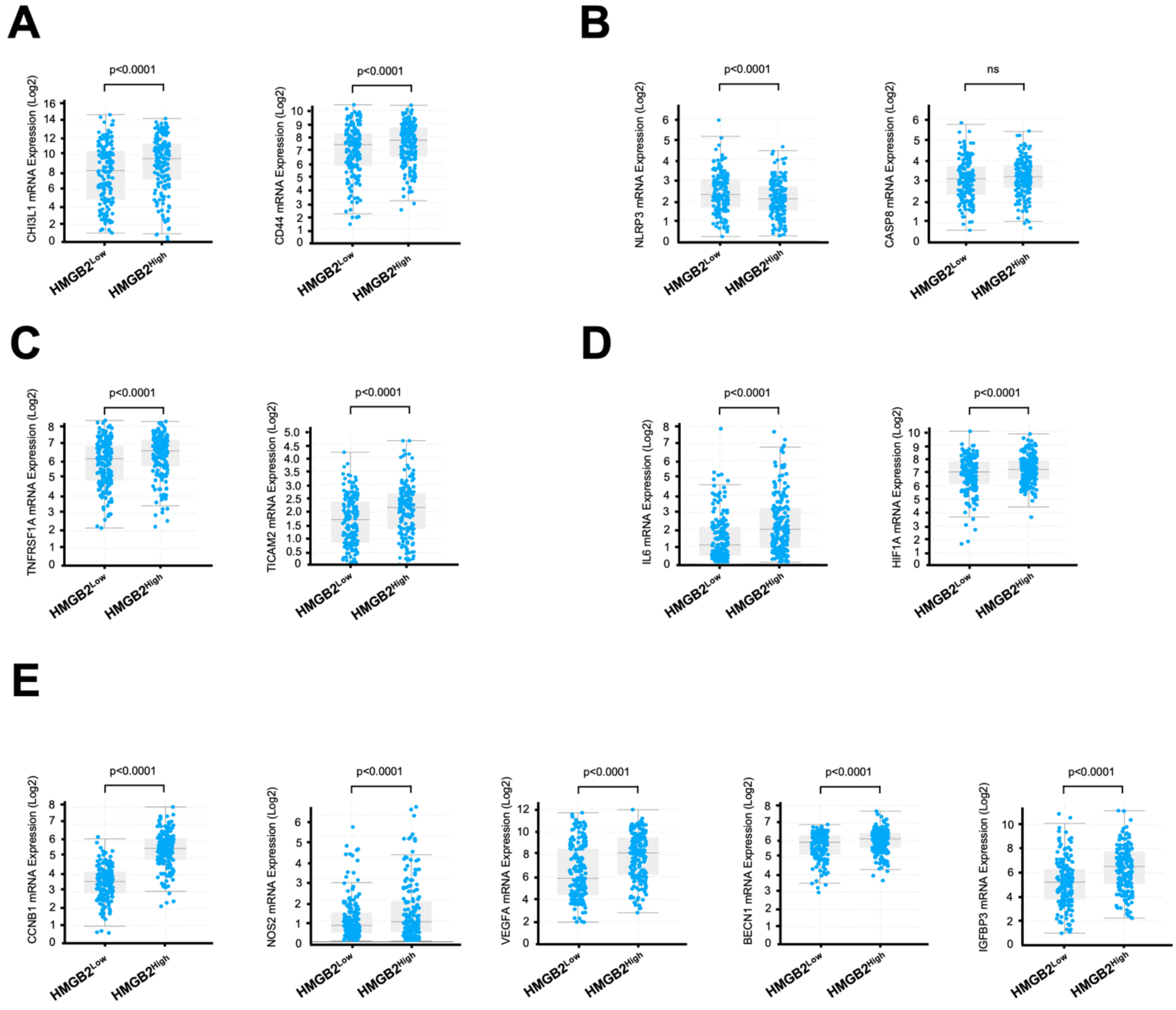


**Figure S19. Additional necrosome associated necroptosis signature in Human glioblastoma patients**. (**A**) Box correlation plot showing mRNA expression (Log2 score) among glioblastoma patients within the GLASS consortium [26] (n=329 patients) for *HMGB2* with MES-cell state (*CHI3L1* and *CD44*) [33]. (**B**) Expression levels with less *NLRP3* transcripts associated with pyroptosis, and no increase in *CASP8*, required for necroptosis activation [34]. (**C**) *TNF* expression and *TICAM2* associated with TLR3/4 activation [34]. (**E**) Correlation plot showing mRNA expression (Log2 score) of inflammatory and hypoxia transcripts of *IL6* and *HIF1A*. (**E**) Box correlation plot showing the terminal glioblastoma aggressive signals of *CCNB1, NOS2, VEGFA, BECN1*, and *IGFBP3* mRNA expression in $HMGB2^{Low}$

*vs. HMGB2*$^{high}$ groups. Two-tailed unpaired t-test were used for statistical comparisons of mRNA correlates.
